# Estimation of Sexual and Gender Minorities in the adult population of Japan: Descriptive Epidemiological Study utilizing a Nationwide Cross-Sectional Internet Survey

**DOI:** 10.1101/2024.06.11.24308803

**Authors:** Tetsuji Minami, Manami Inoue, Midori Matsushima, Takashi Yoshioka, Takahiro Tabuchi

## Abstract

**Background:** Descriptive study of Japanese sexual and gender minority (SGM) population with national representativeness have not been conducted. We sought to estimate the proportion of the Japanese SGM, and to describe those demographic, socioeconomic, and health-related variables.

**Methods:** Utilizing data on a nationwide cross-sectional internet survey from September 12th to October 19th, 2022, we conducted a comprehensive descriptive study by applying inverse probable weighting method for national representativeness. Participants self-reported as heterosexual, homosexual, bisexual, or gender minority (GM) and responded to questions on various demographic, socioeconomic, and health-related concerns.

**Findings:** Among Japanese adults, 4.8% identified as homosexuals, 1.3% as bisexuals, and 3.8% as GMs. SGMs were less likely to be married/partnered compared to heterosexuals, though a certain proportion were in opposite-sex marriages/partnerships. SGMs had lower household equivalent income, insurance coverage, home ownership, current smoking rates, good self-rated health, and full COVID-19 vaccination rates. They also exhibited higher rates of substance use, severe psychological distress, feelings of loneliness, and fear of COVID-19 compared to heterosexuals. When divided by assigned sex at birth, SGM males had poorer employment status, lower academic attainment, and higher body mass index compared to their heterosexual counterparts, while SGM females showed opposite trends.

**Interpretation:** Differences in demographic, socioeconomic, and health status between heterosexuals and SGMs underscore the need for targeted health policies and interventions to address health disparities among Japanese SGMs. Additionally, these results suggest that directly applying Western health policies to the Japanese context may not always be appropriate.

**Funding:** Funded by the Japan Society for the Promotion of Science, the Research Support Program to Apply the Wisdom of the University to tackle COVID-19 Related Emergency Problems, University of Tsukuba, and Health Labour Sciences Research Grantand the Japan Agency for Medical Research and Development.

## Introduction

Sexual and gender minorities (SGMs), commonly known as lesbian, gay, bisexual, and transgender (LGBT) individuals, have been a marginalized population across the globe, often facing discrimination, stigma, and prejudice, leading to health disparities. Minority stress framework theory, which emphasizes the marginalization of SGM, has been widely used to understand the underlying mechanisms of these health disparities.^1^ Various resolutions relating to sexual orientation and gender identity (SOGI) have been endorsed by the United Nations ^2^, and safeguarding the rights of SGM has now become a paramount concern globally. A growing body of SGM research has documented health disparities, such as higher rates of mental health problems, substance use, physical illness, and so on.

The Japanese government, as a member of the United Nations LGBT Core Group ^3^, is actively advocating for the implementation of policy guidelines for safeguarding the rights of SGM in both workplace and educational settings (e.g., laws to prevent LGBT harassment and outing). ^4,5^ Some activists criticized that Japan, as the only G7 nation, has yet to enact legislation prohibiting discrimination against SGM and legalizing same-sex marriage. ^6^ Nevertheless, unlike in the past West, in Japan’s context of philosophic (Confucianism), religious (Shintoism and Buddhism), and cultural norms, homosexuality had been neither criminalized, tabooed nor stigmatized, but rather partially accepted as high-society culture (*shud*ō, *kagema*, and so on) and popular art form (*yaoi*, *yuri*, and so on). ^7^ If the familial norms of marriage and procreation are adhered to, the diversity of SOGI is tolerated, and SGM sometimes enjoys same-sex relationships with social confidentiality while being married to the opposite sex. ^7,8^ Furthermore, Japan reportedly had a lower level of both country-level structural stigma for SGM and the concealment of SOGI among Asian countries.^9^

To the best of our knowledge, there are no Asian or Japanese studies regarding SGM with national representativeness. A few Asian studies evaluating specific cohorts have reported that SGM is marginalized ^10–12^, whereas much of the literature has been conducted in Western countries. The paucity of evidence could be attributed to certain Asian countries’ opposition to the aforementioned United Nations resolution regarding SGM. ^2^ Furthermore, criminalization and penalty still exist regarding movements and expressions affiliated with SGM and the prohibition and penalization of same-sex sexual acts in certain Asian countries. ^13^ Some studies of SGM Asian Americans revealed their increased marginalization relative to both SGM and heterosexual ethnic majorities; nevertheless, such an effect was modified by the intersectionality of racism and heterosexism. ^8,14^ Previous studies give it little generalizability to Asian countries.

In Japan, as an industrialized country with a unique culture that differed from the West, ascertaining the proportion of SGM, and evaluating demographic characteristics, household relationships, and health outcomes would provide essential information and insight to policymakers, healthcare professionals, and researchers, contributing to developing appropriate interventions to improve the health disparities if exists. Furthermore, filling the knowledge gap between Western countries and Japan would advance establishing international policies to safeguard SGM’s rights, freedom, and dignity. Here, we aimed to estimate the proportion of the Japanese SGM population and describe those demographic characteristics using nationwide internet-based survey data.

## Materials and methods

### Data collection, study participants, and exclusion

This study used data from the 3rd wave of the COVID-19 and Society Internet Survey in Japan (JACSIS 2022), conducted by Rakuten Insight Inc. ^15^ from September 12 to October 19, 2022. Participants were recruited from a panel of 2.3 million Japanese individuals, ensuring diverse representation. They provided demographic information upon registration and consented to participate, with minors providing parental consent. The initial JACSIS study used multistage sampling, stratifying by age and sex, and reached a sample size of 28,000 through email invitations. For JACSIS 2022, the target sample size was 32,000, with priority invitations sent to previous respondents, achieving a 65.7% response rate (27,327 respondents).

Two algorithms excluded ineligible respondents. Initially, 3,370 were excluded based on unrealistic household sizes, incorrect responses to a directed question, and straight-line responses on comorbidities and substance use. Subsequently, 592 were excluded for reporting non-binary sex assigned at birth for themselves or their partners. The final sample consisted of 28,038 respondents. Detailed methodology of the JACSIS study series is in previous studies, with further details in the **Supplementary Methods**. A summary flowchart of participants is shown in **Figure 1**.

**Figure 1.**
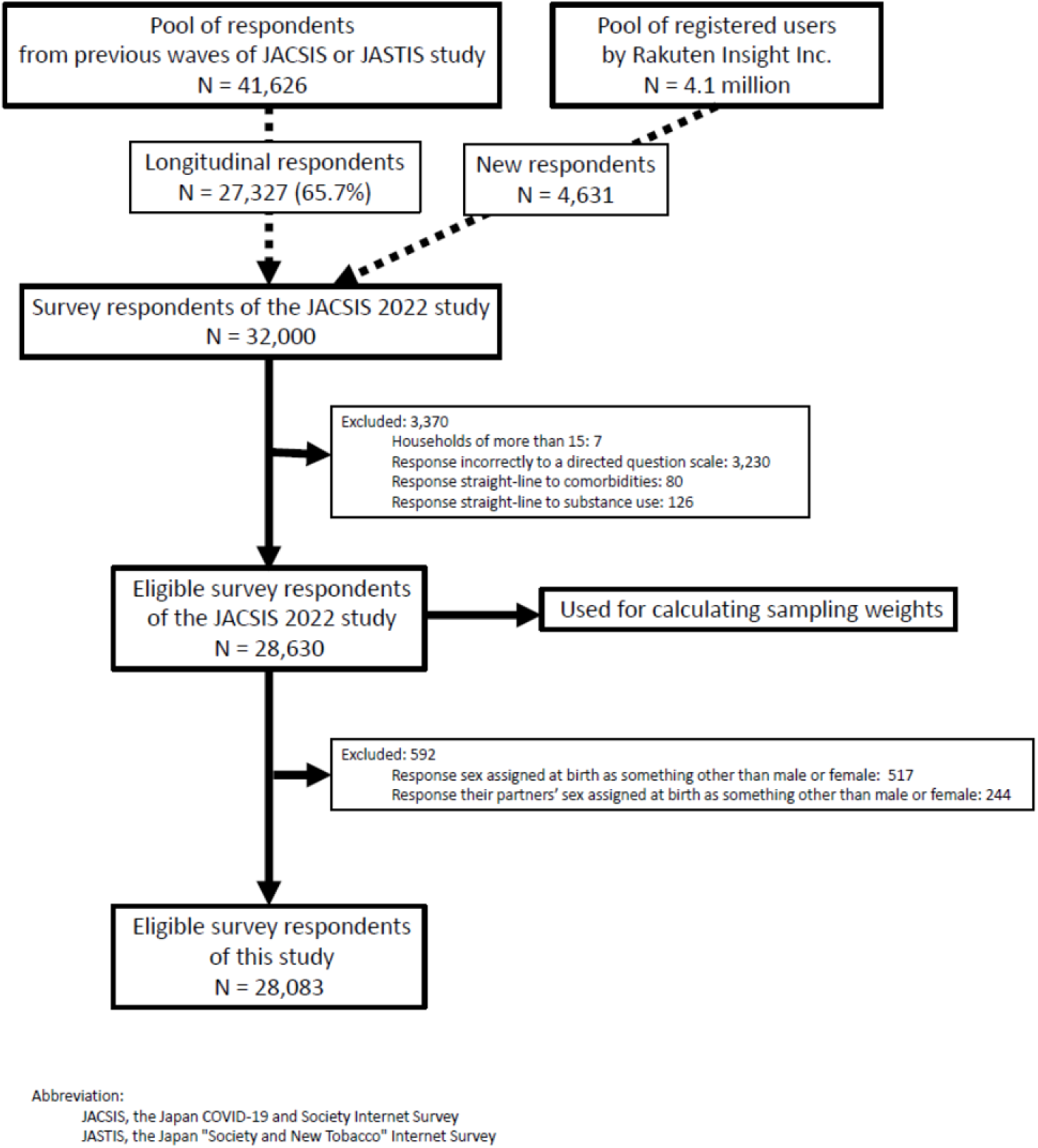
Flowchart for eligible study participants.

### Ascertainment of sexual orientation and gender identity

**Table 1** presents a survey questionnaire to determine SOGI. The questionnaire design followed a “two-step approach” frequently employed in previous research ^16^. Due to the agency’s programmatic setup of the survey, each sub-question within a single main question was assigned the same response items, causing inappropriate response items for the respondents’ and their partners’ assigned sex at birth. As per the “two-step approach”, respondents should answer with “male” or “female” for these sub-questions. The sentence “Choose either 1 (male) or 2 (female)” was added following these sub-questions to serve as a directed question scale for the exclusion criteria, as previously mentioned.

**Table 1.**
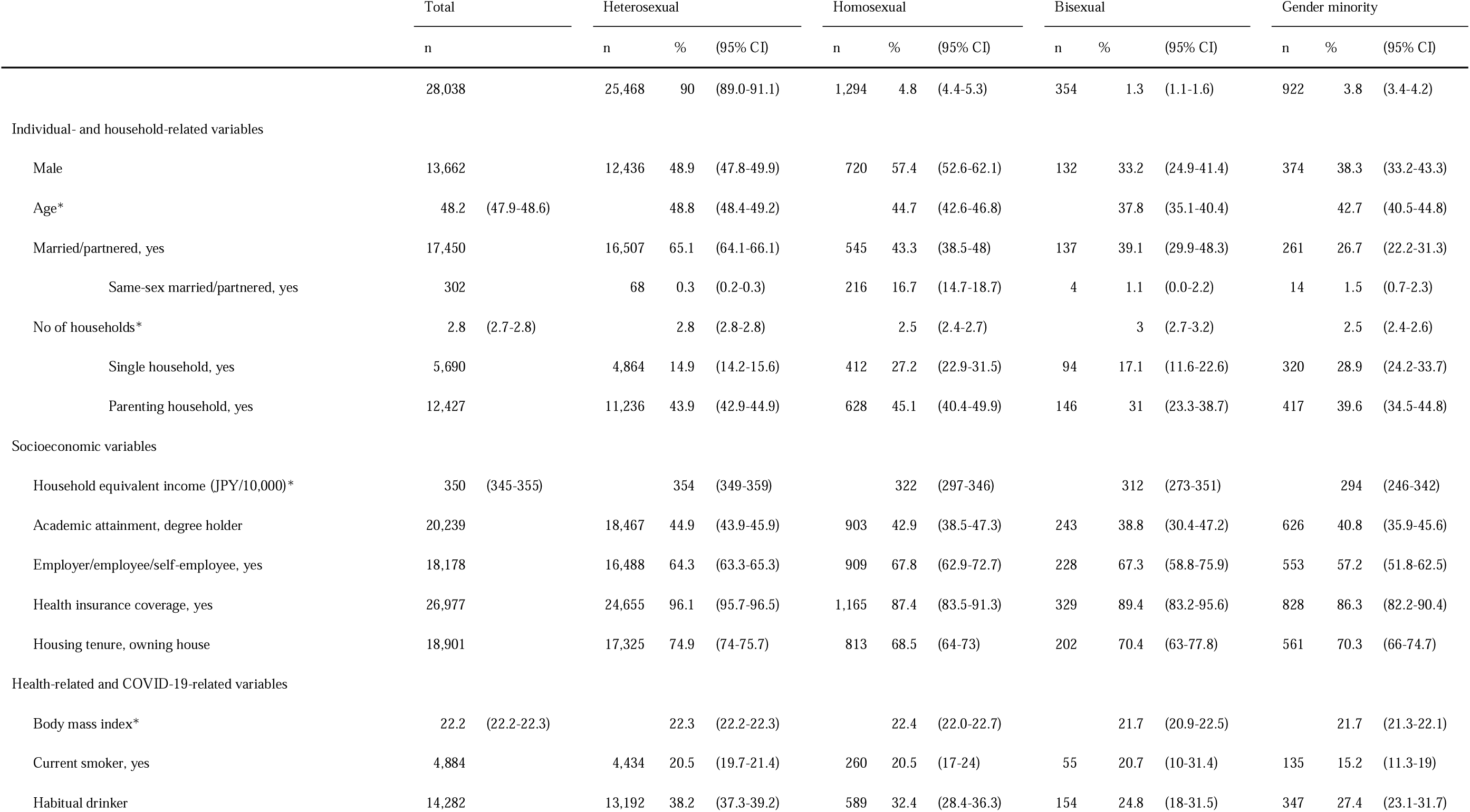
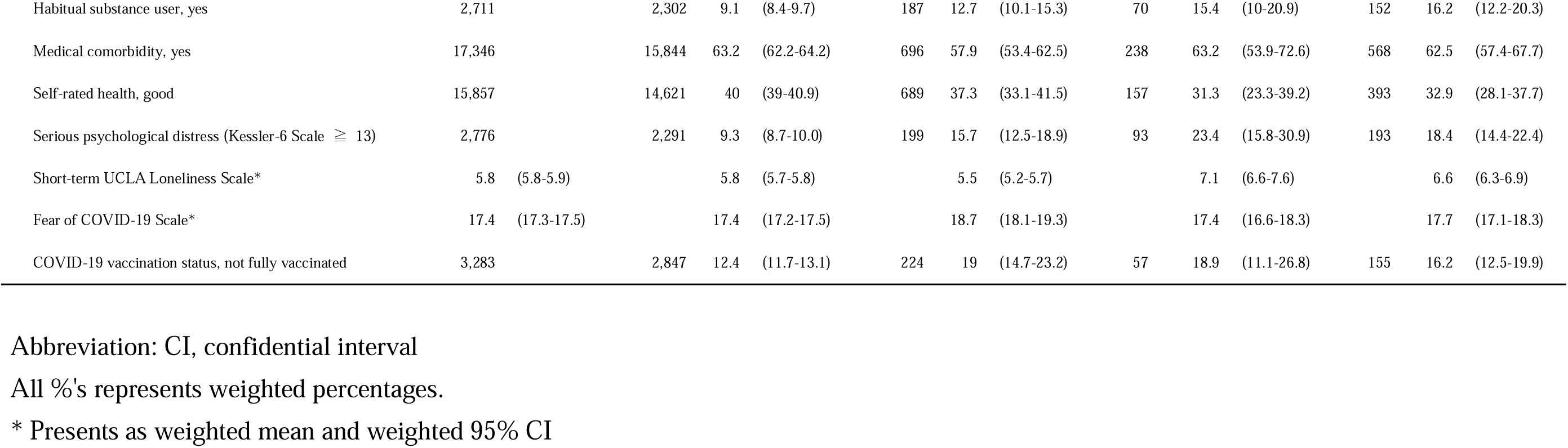
Summary characteristics of respondents by sexual orientation and gender identity.

Initially, individuals whose sex assigned at birth did not match their gender identity were categorized into “gender minority” (GM), such as those who identified as transgender, questioning, or asexual. Subsequently, respondents whose gender identity aligned with their assigned sex at birth were categorized based on their sexual orientation as “heterosexual” “homosexual” or “bisexual”.

### Variables measure

The JACSIS 2022 survey consists of 92 main questionnaires. To ensure complete responses, incentives were only provided to participants who answered all questions, resulting in no missing values in the collected data. The variables utilized in this descriptive epidemiological study are as follows: age, marital status, household components and composition, household equivalent income, academic attainment, employment status, housing tenure, health insurance coverage, housing tenure, body mass index (BMI), smoking status, alcohol consumption, substance use, medical comorbidities, self-rated health, psychological distress measured by the Kessler-6 Scale, Short-term UCLA Loneliness Scale Japanese version, the Fear of COVID-19 Scale, and COVID-19 vaccination status. The precise questionnaire and precise categorization of each variable are given in the **Supplementary Methods**.

### Study approval

The research ethics committees of the Osaka International Cancer Institute and National Cancer Center approved the study protocol (approval number: 20084 and 2020-447, respectively).

### Statistical analyses

Sampling weights for this study were derived from nationally representative samples of Comprehensive Survey of Living Conditions of People on Health and Welfare 2019 (CSLCPHW) obtained from the Ministry of Health, Labour, and Welfare. JACSIS 2022 and CSLCPHW 2019 data were merged and used in a logistic regression model to estimate propensity scores. The sampling weights, calculated for 28,630 respondents after initial exclusions, were predicted from a logistic model adjusting for several socio-economic variables using CSLCPHW 2019 data. The outcome variable in the logistic model was the type of survey (JACSIS 2022 or CSLCPHW 2019), and the sampling weight represented the probability of “being an internet survey respondent.”

An inverse probability weighting method was applied to adjust for potential sampling bias ^17^. Descriptive analyses included unweighted numbers and weighted proportions (henceforth, written as “proportion”) with 95% confidence intervals (CIs) for discrete variables and weighted means with 95% CIs for continuous variables, calculated using a robust variance estimator to account for inverse probability weighting. Analyses by sex assigned at birth and SOGI categories were also performed. All analyses were conducted using STATA version 17 (Stata Corp).

## Results

Among Japanese adults, 4.8% identified as homosexual, 1.3% as bisexual, and 3.8% as GM. There were more homosexuals and fewer bisexuals and GMs among males than females.

SGMs were more common in younger age groups under age 30, although there was no age-related variability in the proportion of SGM after age 30. SGM females were more common in all age groups over 20 compared to males (**Table 2**, Supplementary Table 1, & Figure 2).

**Figure 2.**
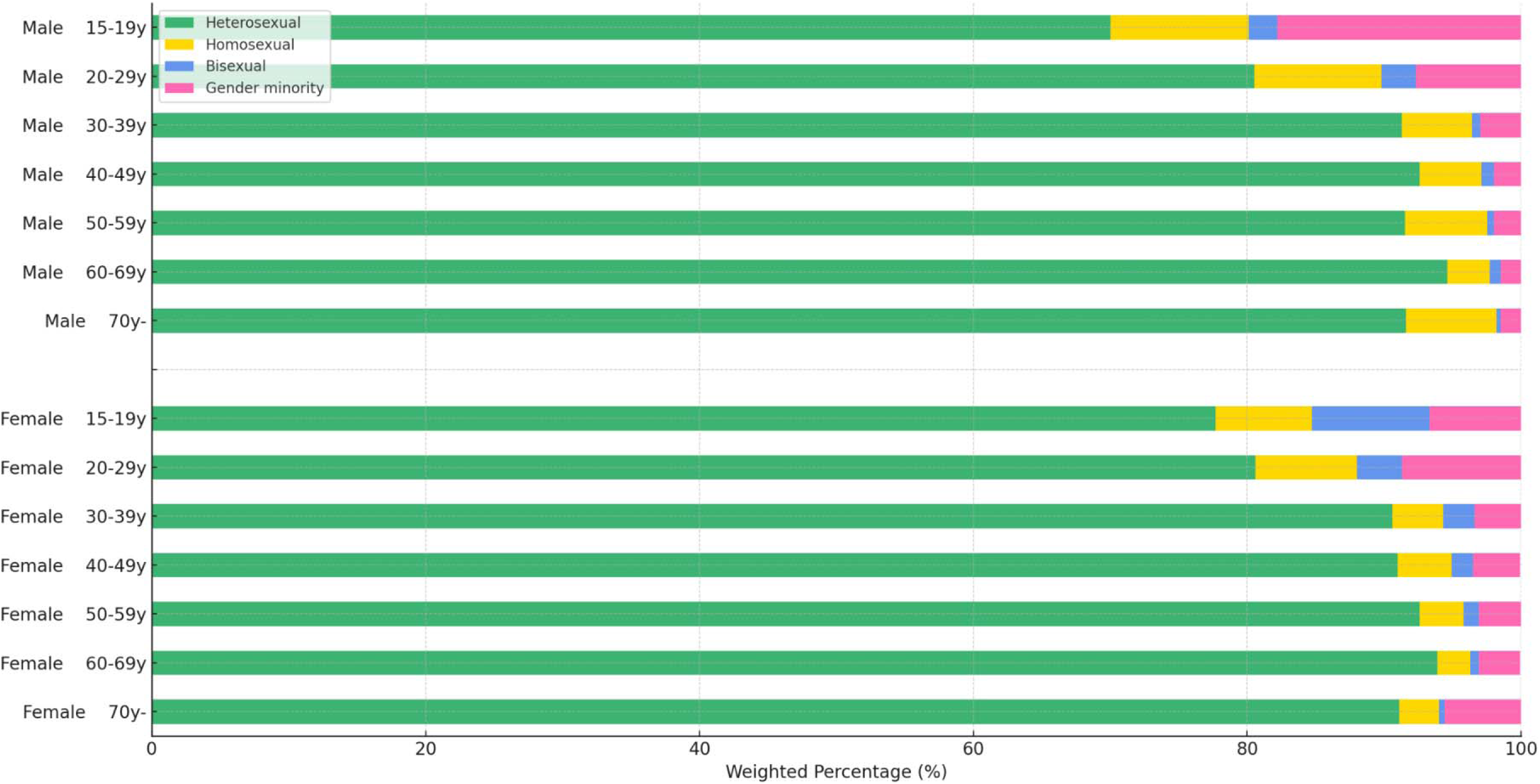
Sexual Orientation and Gender Identity Categories According to Age Group and Sex.

**Table 2.** SOGI category by age group and sex.

|  |  | Total | Heterosexual |  |  |  | Homosexual |  |  | Bisexual |  |  | Gender minority |  |
| --- | --- | --- | --- | --- | --- | --- | --- | --- | --- | --- | --- | --- | --- | --- |
| Assigned sex at birth |  |  |  |  |  |  |  |  |  |  |  |  |  |  |
| Male |  | 13,662 |  |  |  |  |  |  |  |  |  |  |  |  |
| 15-19 | n, %, (95%CI) | 83 | 65 | 70.0 | (44.6-95.4) | 8 | 10.1 | (-0.6-20.9) | 4 | 2.1 | (-0.3-4.5) | 6 | 17.8 | (-9.1-44.6) |
| 20-29 | n, %, (95%CI) | 1,932 | 1,605 | 80.5 | (78.2-82.8) | 168 | 9.3 | (7-11.6) | 42 | 2.5 | (1.7-3.3) | 117 | 7.7 | (5.6-9.8) |
| 30-39 | n, %, (95%CI) | 3,098 | 2,820 | 91.3 | (89.7-92.9) | 152 | 5.1 | (4-6.3) | 29 | 0.6 | (0.3-0.8) | 97 | 3 | (2.1-3.9) |
| 40-49 | n, %, (95%CI) | 2,318 | 2,141 | 92.6 | (91-94.2) | 101 | 4.5 | (3.3-5.7) | 25 | 0.9 | (0.5-1.4) | 51 | 2 | (1.4-2.5) |
| 50-59 | n, %, (95%CI) | 2,053 | 1,870 | 91.5 | (89.8-93.1) | 127 | 6.0 | (4.8-7.2) | 12 | 0.5 | (0.1-1) | 44 | 2 | (1.2-2.8) |
| 60-69 | n, %, (95%CI) | 2,065 | 1,949 | 94.6 | (93.2-95.9) | 76 | 3.1 | (2.1-4.2) | 14 | 0.8 | (0.2-1.5) | 26 | 1.5 | (0.7-2.2) |
| 70- | n, %, (95%CI) | 2,113 | 1,986 | 91.6 | (88.8-94.4) | 88 | 6.6 | (3.8-9.3) | 6 | 0.3 | (0-0.6) | 33 | 1.6 | (0.9-2.2) |
| Female |  | 14,376 |  |  |  |  |  |  |  |  |  |  |  |  |
| 15-19 | n, %, (95%CI) | 130 | 95 | 77.7 | (63.3-92.1) | 12 | 7.0 | (1.5-12.4) | 9 | 8.6 | (-1.3-18.6) | 14 | 6.7 | (-0.1-13.5) |
| 20-29 | n, %, (95%CI) | 2,442 | 2,016 | 80.6 | (77.9-83.2) | 185 | 7.4 | (5.8-9) | 89 | 3.3 | (1.9-4.8) | 152 | 8.7 | (6.5-10.9) |
| 30-39 | n, %, (95%CI) | 3,077 | 2,857 | 90.6 | (88.2-93) | 94 | 3.7 | (1.8-5.6) | 45 | 2.3 | (0.9-3.8) | 81 | 3.4 | (2.2-4.5) |
| 40-49 | n, %, (95%CI) | 2,356 | 2,161 | 91.0 | (88.8-93.3) | 81 | 3.9 | (2.5-5.4) | 33 | 1.6 | (0.7-2.5) | 81 | 3.4 | (2-4.8) |
| 50-59 | n, %, (95%CI) | 2,044 | 1,878 | 92.6 | (90.6-94.6) | 65 | 3.2 | (2.2-4.1) | 21 | 1.1 | (0.3-2) | 80 | 3.1 | (1.5-4.7) |
| 60-69 | n, %, (95%CI) | 2,155 | 2,020 | 93.9 | (91.7-96.2) | 62 | 2.4 | (1.2-3.6) | 15 | 0.6 | (0-1.3) | 58 | 3 | (1.6-4.3) |
| 70- | n, %, (95%CI) | 2,172 | 2,005 | 91.1 | (89-93.1) | 75 | 2.9 | (1.6-4.2) | 10 | 0.4 | (-0.1-0.8) | 82 | 5.6 | (4.1-7.2) |

Abbreviation: SOGI, sexual orientation and gender identity; CI, confidential interval
All %'s represents weighted percentages.

**Table 3.** Same-sex marriages/partnership among married/partnered respondents by SOGI category and sex.

|  | Heterosexual (N=16507) |  |  | Homosexual (N=545) |  |  | Bisexual (N=137) |  |  | Gender minority (N=261) |  |  |
| --- | --- | --- | --- | --- | --- | --- | --- | --- | --- | --- | --- | --- |
|  | n/total* | % | (95% CI) | n/total* | % | (95% CI) | n/total* | % | (95% CI) | n/total* | % | (95% CI) |
| Assigned sex at birth |  |  |  |  |  |  |  |  |  |  |  |  |
| Male | 29/7950 | 0.4 | (0.2-0.6) | 128/287 | 56.2 | (46.7-65.7) | 4/44 | 6.7 | (-0.7-14.2) | 10/106 | 7.2 | (1.8-12.6) |
| aged 15-49 years | 18/3406 | 0.7 | (0.3-1.1) | 60/104 | 64.4 | (51.0-77.8) | 4/23 | 14.3 | (-1.1-29.8) | 7/53 | 7.0 | (1.3-12.7) |
| aged over 50 years | 11/4544 | 0.2 | (0.0-0.4) | 68/183 | 51.1 | (37.8-64.4) | 0/21 | 0.0 |  | 3/53 | 7.4 | (-1.8-16.5) |
| Female | 39/8557 | 0.3 | (0.2-0.5) | 88/258 | 32.6 | (22.9-42.3) | 0/93 | 0.0 |  | 4/155 | 0.7 | (-0.1-1.6) |
| aged 15-49 years | 20/4541 | 0.4 | (0.2-0.7) | 51/125 | 42.2 | (27.3-57.2) | 0/67 | 0.0 |  | 2/63 | 1.0 | (-0.6-2.6) |
| aged over 50 years | 19/4016 | 0.2 | (0.1-0.4) | 35/128 | 21.9 | (12.5-31.3) | 0/23 | 0.0 |  | 2/89 | 0.6 | (-0.2-1.4) |

Abbreviation: SOGI, sexual orientation and gender identity; CI, confidential interval
All %'s represents weighted percentages.
\* Presents as total of married/partnered respondents

**Table 4.** Parenting household among married/partnered respondents by SOGI category and sex.

|  |  | Heterosexual |  |  | Homosexual |  |  | Bisexual |  |  | Gender minority |  |  |
| --- | --- | --- | --- | --- | --- | --- | --- | --- | --- | --- | --- | --- | --- |
|  |  | n/total* | % | (95% CI) | n/total* | % | (95% CI) | n/total | % | (95% CI) | n/total | % | (95% CI) |
| Assigned sex at birth |  |  |  |  |  |  |  |  |  |  |  |  |  |
| Male |  |  | 43.9 | (42.2-45.7) |  | 40.6 | (31.2-50.1) |  | 30.7 | (12.6-48.9) |  | 32.7 | (19.4-46.0) |
|  | Same-sex marriage | 13/29 | 45.1 | (19.9-70.2) | 56/128 | 41.5 | (27.2-55.8) | 1/4 | 29.5 | (-20.2-79.2) | 6/10 | 56.9 | (18.1-95.6) |
|  | Opposite-sex marriage | 3237/7921 | 43.9 | (42.2-45.7) | 59/159 | 39.5 | (27.6-51.3) | 15/40 | 30.8 | (11.7-50.0) | 27/95 | 30.8 | (16.8-44.9) |
| Female |  |  | 37.4 | (35.7-39.2) |  | 34.8 | (24.4-45.2) |  | 30.5 | (15.3-45.7) |  | 28.1 | (16.8-39.4) |
|  | Same-sex marriage | 7/39 | 29.0 | (4.5-53.5) | 28/86 | 42.5 | (23.7-61.4) | 0 |  |  | 1/4 | 11.2 | (-12.0-34.3) |
|  | Opposite-sex marriage | 2857/8518 | 37.5 | (35.7-39.2) | 54/170 | 31 | (19.1-43.0) | 27/93 | 30.5 | (15.3-45.7) | 44/151 | 28.3 | (16.9-39.6) |

Abbreviation: SOGI, sexual orientation and gender identity; CI, confidential interval
5 All %'s represents weighted percentages.

**Table 5.**
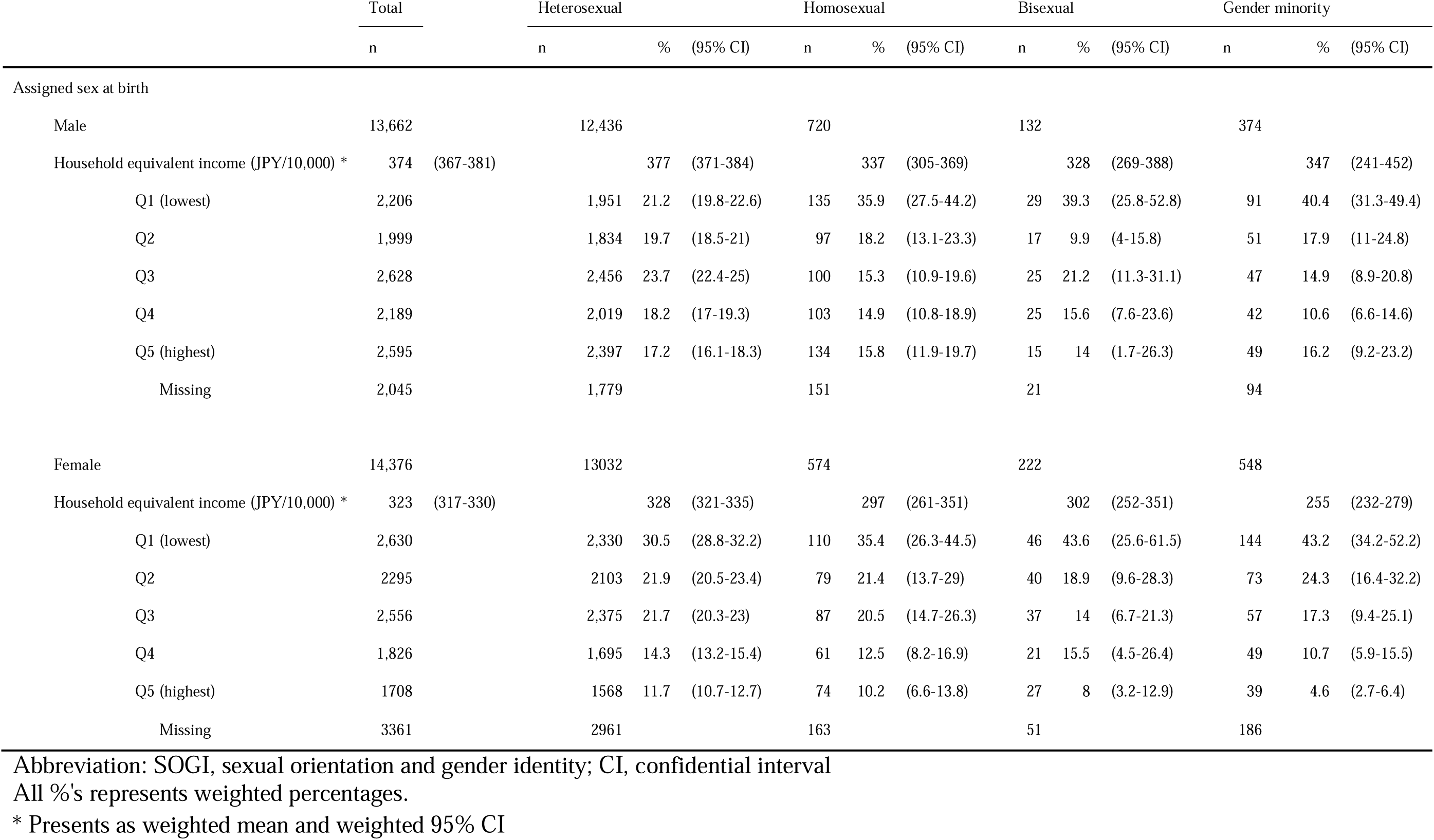
Household equivalent income by SOGI category and sex.

**Table 6.**
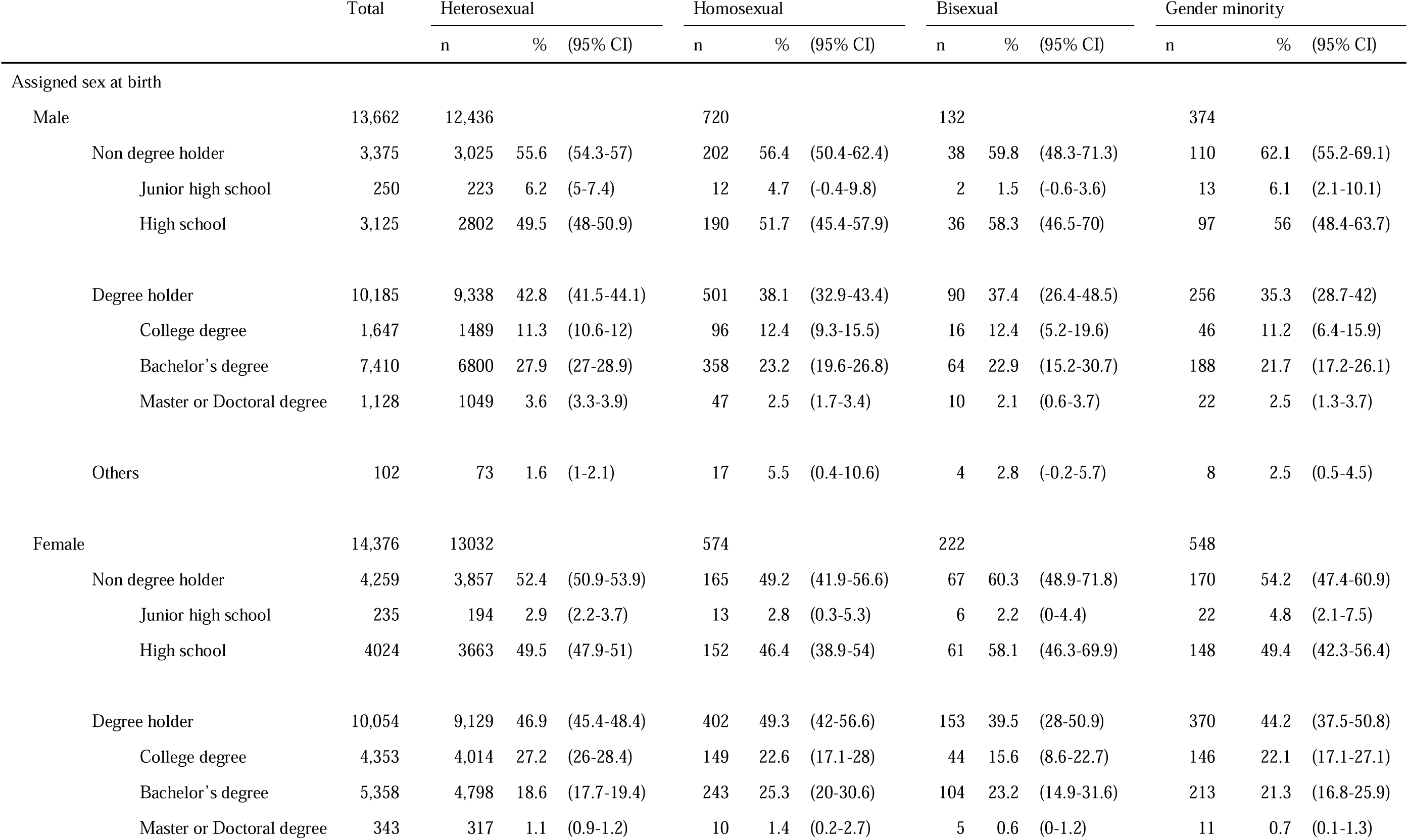
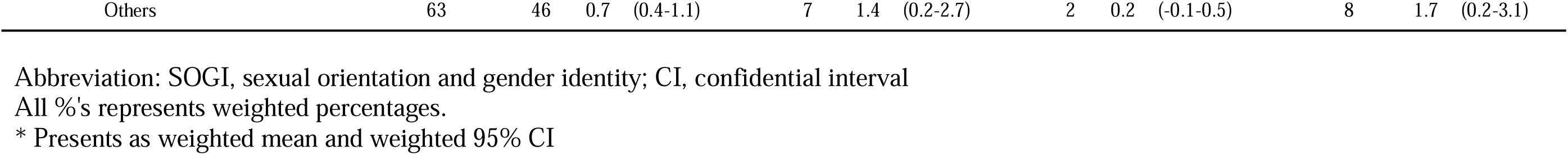
Academic attainment by SOGI category and sex.

**Table 7.**
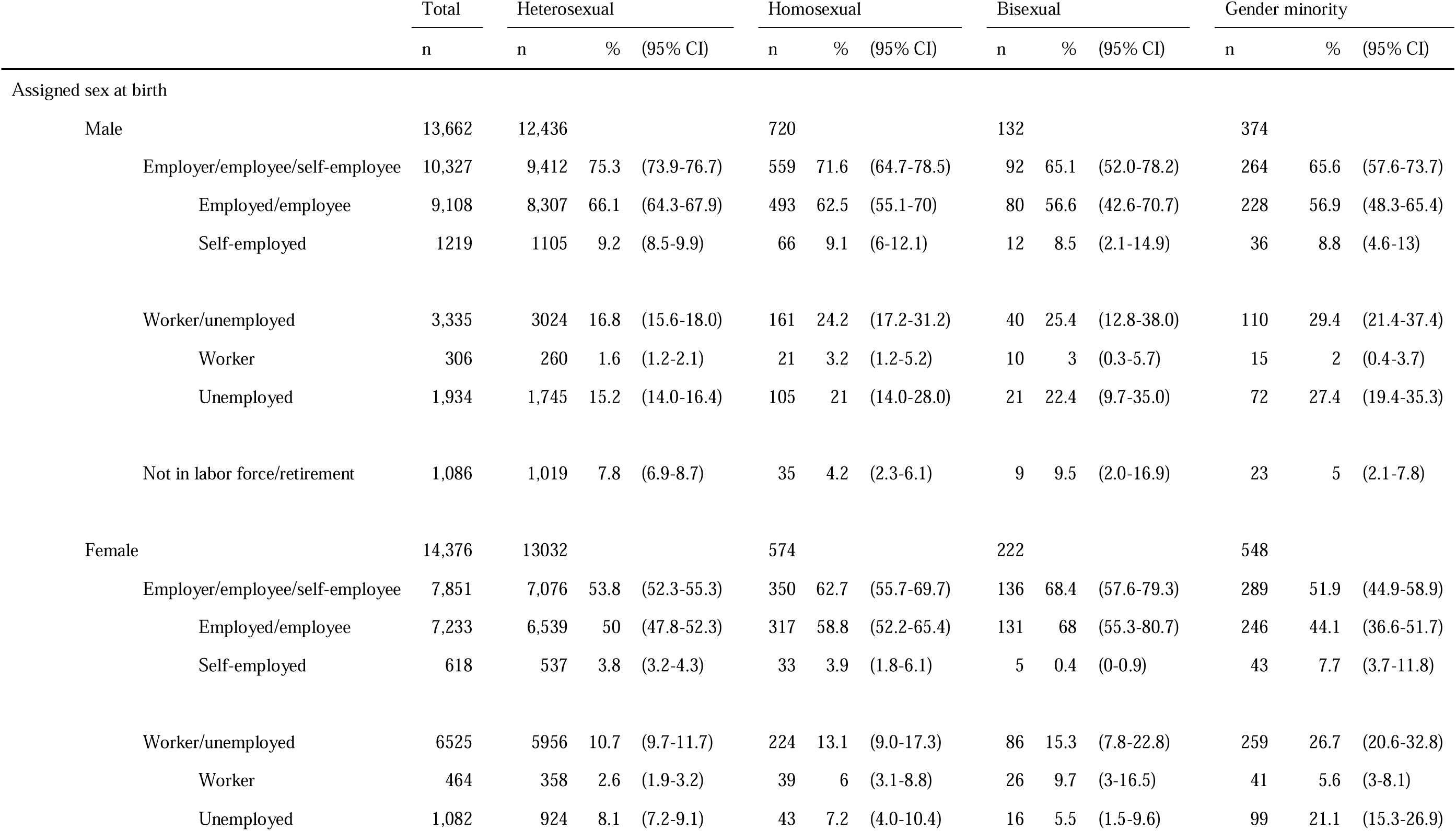
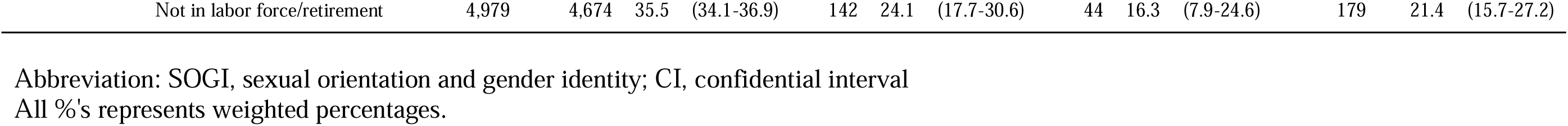
Employment status by SOGI category and sex.

**Table 8.** Health insurance coverage by SOGI category and sex.

|  | Total | Heterosexual |  |  | Homosexual |  |  | Bisexual |  |  | Gender minority |  |  |
| --- | --- | --- | --- | --- | --- | --- | --- | --- | --- | --- | --- | --- | --- |
|  | n | n | % | (95% CI) | n | % | (95% CI) | n | % | (95% CI) | n | % | (95% CI) |
| Assigned sex at birth |  |  |  |  |  |  |  |  |  |  |  |  |  |
| Male | 13,662 | 12,436 |  |  | 720 |  |  | 132 |  |  | 374 |  |  |
| Health insurance coverage, yes |  |  | 95.9 | (95.3-96.5) |  | 86.8 | (81.4-92.3) |  | 83.9 | (71.3-96.5) |  | 84.9 | (79.4-90.5) |
| Employer-based health insurance | 7,275 | 6704 | 51.9 | (50.4-53.3) | 328 | 39.9 | (34.2-45.7) | 77 | 56 | (42.9-69.1) | 166 | 43.1 | (34.9-51.4) |
| Residence-based National health insurance | 5,823 | 5309 | 44.1 | (42.6-45.5) | 315 | 46.9 | (40.6-53.2) | 44 | 27.9 | (17.7-38.2) | 155 | 41.8 | (34-49.6) |
| Health insurance coverage, no |  |  | 4.1 | (3.5-4.7) |  | 13.2 | (7.7-18.6) |  | 16.1 | (3.5-28.7) |  | 15.1 | (9.5-20.6) |
| Welfare | 62 | 43 | 0.5 | (0.2-0.8) | 9 | 1.1 | (0.3-1.9) | 2 | 1.9 | (-1.3-5) | 8 | 1.9 | (0.1-3.7) |
| No insurance | 137 | 106 | 0.9 | (0.6-1.2) | 20 | 3 | (1.2-4.8) | 2 | 5.8 | (-3.5-15.1) | 9 | 3.1 | (-0.1-6.4) |
| Others | 365 | 274 | 2.6 | (2.2-3.1) | 48 | 9.1 | (3.8-14.4) | 7 | 8.4 | (-1.1-17.9) | 36 | 10 | (5.6-14.4) |
| Female | 14,376 | 13032 |  |  | 574 |  |  | 222 |  |  | 548 |  |  |
| Health insurance coverage, yes |  |  | 96.2 | (95.6-96.8) |  | 88.2 | (82.6-93.8) |  | 92.2 | (85.6-98.7) |  | 87.1 | (81.4-92.9) |
| Employer-based health insurance | 7,933 | 7,297 | 54.8 | (53.3-56.3) | 274 | 49.9 | (42.6-57.3) | 124 | 53.7 | (40.8-66.6) | 238 | 37.4 | (30.8-43.9) |
| Residence-based National health insurance | 5,946 | 5,345 | 41.4 | (39.9-43) | 248 | 38.3 | (31.2-45.3) | 84 | 38.4 | (26.2-50.6) | 269 | 49.8 | (42.7-56.8) |
| Health insurance coverage, no |  |  | 3.8 | (3.2-4.4) |  | 11.8 | (6.2-17.4) |  | 7.8 | (1.3-14.4) |  | 12.9 | (7.1-18.6) |
| Welfare | 54 | 42 | 0.4 | (0.2-0.6) | 3 | 0.6 | (-0.3-1.5) | 4 | 2.8 | (-1.1-6.8) | 5 | 1.8 | (-1.3-4.8) |
| No insurance | 116 | 84 | 0.7 | (0.5-1) | 14 | 2.7 | (0.8-4.7) | 5 | 4.4 | (-0.9-9.7) | 13 | 3.2 | (0.9-5.4) |
| Others | 327 | 264 | 2.7 | (2.2-3.2) | 35 | 8.5 | (3.1-13.8) | 5 | 0.6 | (0-1.2) | 23 | 7.9 | (3.2-12.7) |

Abbreviation: SOGI, sexual orientation and gender identity; CI, confidential interval
All %'s represents weighted percentages.

**Table 9.**
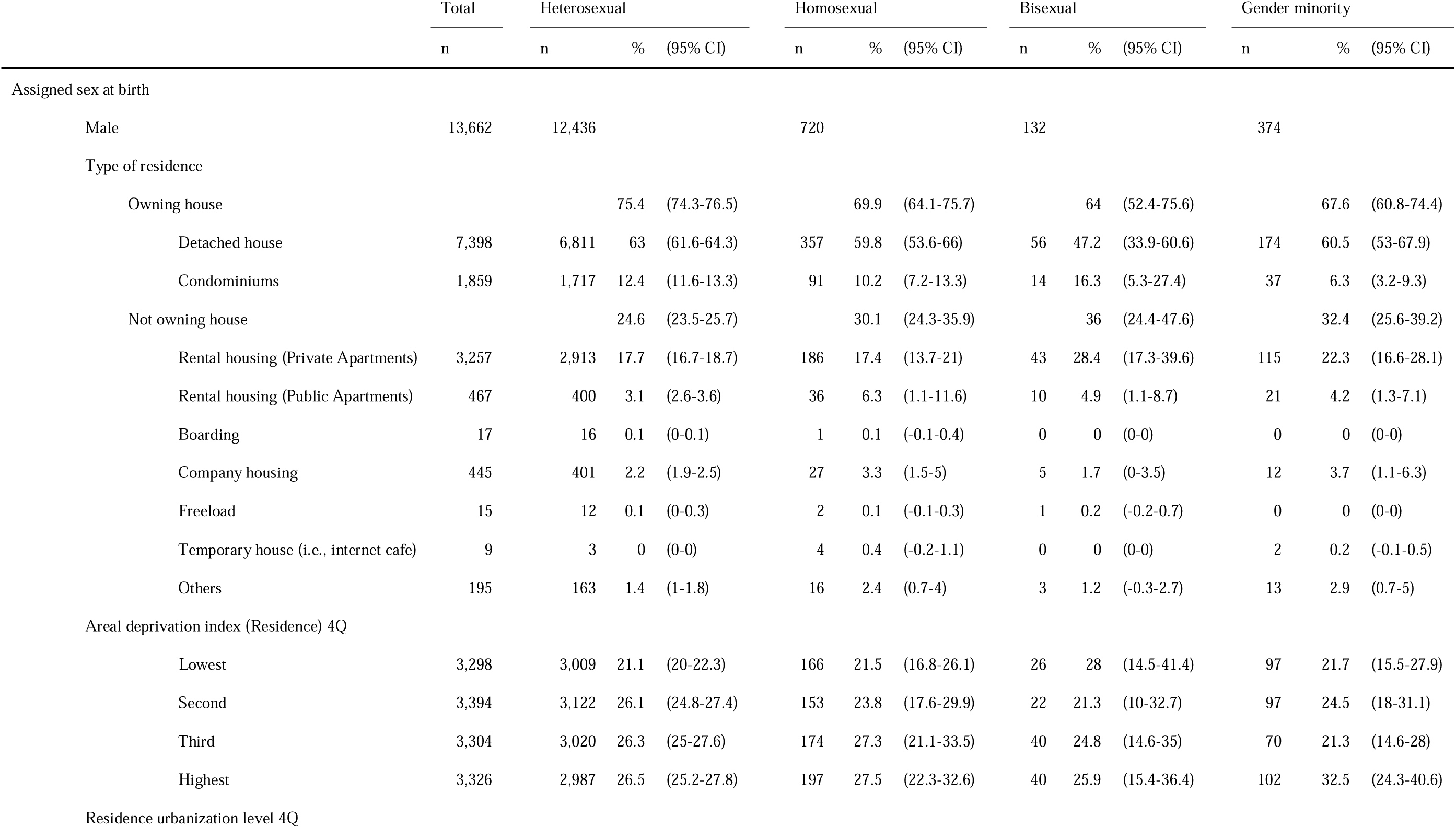
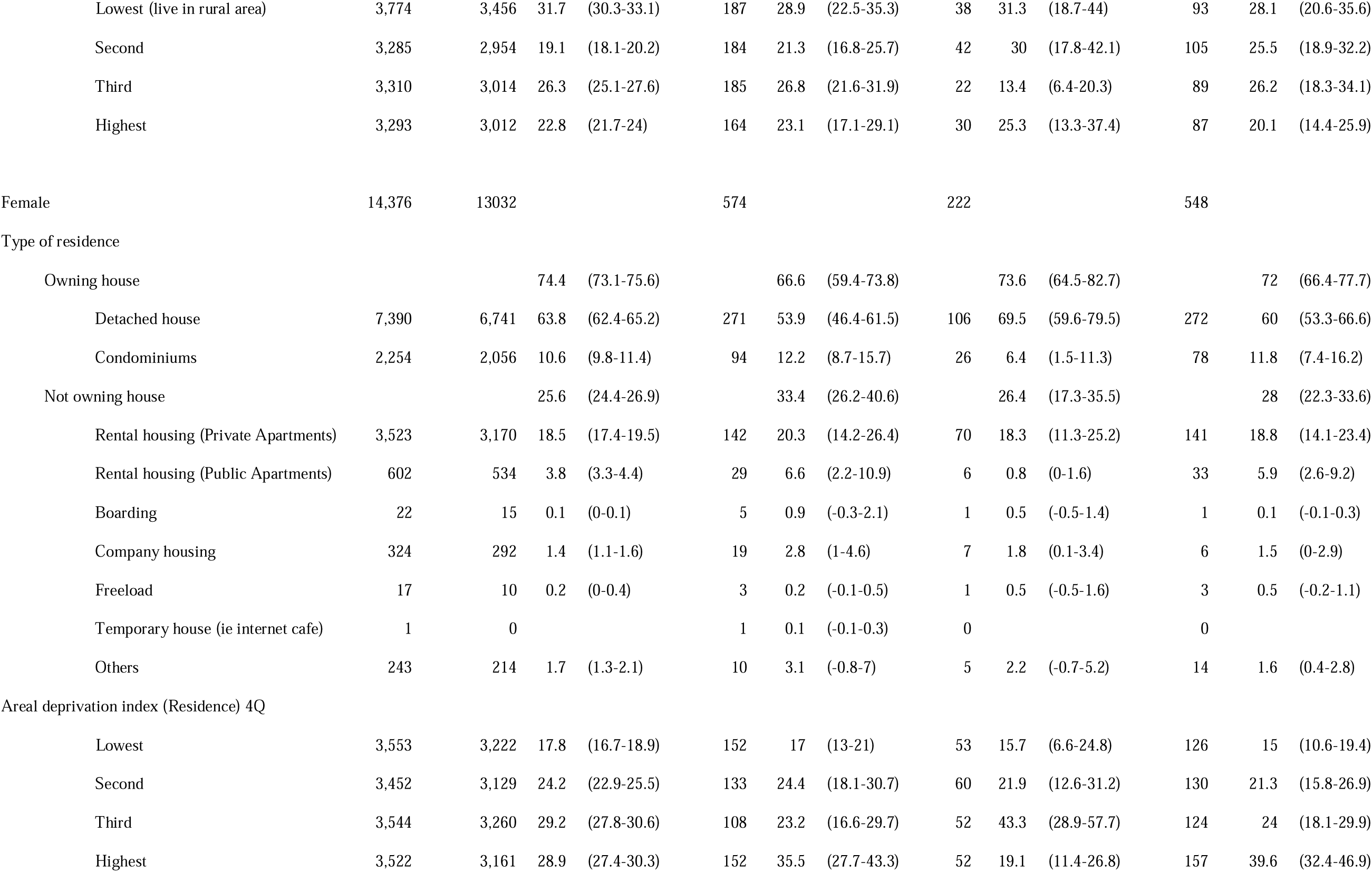
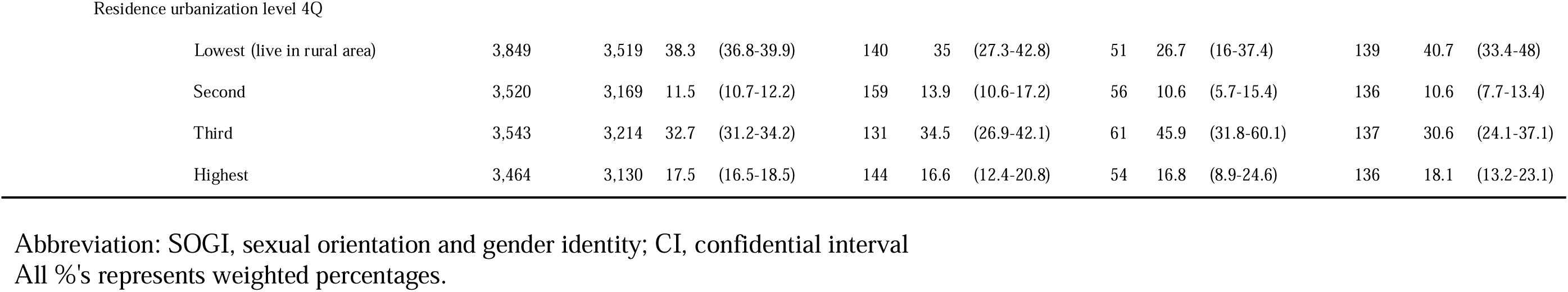
Housing tenure by SOGI category and sex.

|  | Total | Heterosexual |  |  | Homosexual |  |  | Bisexual |  |  | Gender minority |  |  |
| --- | --- | --- | --- | --- | --- | --- | --- | --- | --- | --- | --- | --- | --- |
|  | n | n | % | (95% CI) | n | % | (95% CI) | n | % | (95% CI) | n | % | (95% CI) |
| Assigned sex at birth |  |  |  |  |  |  |  |  |  |  |  |  |  |
| Male | 13,662 | 12,436 |  |  | 720 |  |  | 132 |  |  | 374 |  |  |
| Type of residence |  |  |  |  |  |  |  |  |  |  |  |  |  |
| Owning house |  |  | 75.4 | (74.3-76.5) |  | 69.9 | (64.1-75.7) |  | 64 | (52.4-75.6) |  | 67.6 | (60.8-74.4) |
| Detached house | 7,398 | 6,811 | 63 | (61.6-64.3) | 357 | 59.8 | (53.6-66) | 56 | 47.2 | (33.9-60.6) | 174 | 60.5 | (53-67.9) |
| Condominiums | 1,859 | 1,717 | 12.4 | (11.6-13.3) | 91 | 10.2 | (7.2-13.3) | 14 | 16.3 | (5.3-27.4) | 37 | 6.3 | (3.2-9.3) |
| Not owning house |  |  | 24.6 | (23.5-25.7) |  | 30.1 | (24.3-35.9) |  | 36 | (24.4-47.6) |  | 32.4 | (25.6-39.2) |
| Rental housing (Private Apartments) | 3,257 | 2,913 | 17.7 | (16.7-18.7) | 186 | 17.4 | (13.7-21) | 43 | 28.4 | (17.3-39.6) | 115 | 22.3 | (16.6-28.1) |
| Rental housing (Public Apartments) | 467 | 400 | 3.1 | (2.6-3.6) | 36 | 6.3 | (1.1-11.6) | 10 | 4.9 | (1.1-8.7) | 21 | 4.2 | (1.3-7.1) |
| Boarding | 17 | 16 | 0.1 | (0-0.1) | 1 | 0.1 | (-0.1-0.4) | 0 | 0 | (0-0) | 0 | 0 | (0-0) |
| Company housing | 445 | 401 | 2.2 | (1.9-2.5) | 27 | 3.3 | (1.5-5) | 5 | 1.7 | (0-3.5) | 12 | 3.7 | (1.1-6.3) |
| Freeload | 15 | 12 | 0.1 | (0-0.3) | 2 | 0.1 | (-0.1-0.3) | 1 | 0.2 | (-0.2-0.7) | 0 | 0 | (0-0) |
| Temporary house (i.e., internet cafe) | 9 | 3 | 0 | (0-0) | 4 | 0.4 | (-0.2-1.1) | 0 | 0 | (0-0) | 2 | 0.2 | (-0.1-0.5) |
| Others | 195 | 163 | 1.4 | (1-1.8) | 16 | 2.4 | (0.7-4) | 3 | 1.2 | (-0.3-2.7) | 13 | 2.9 | (0.7-5) |
| Areal deprivation index (Residence) 4Q |  |  |  |  |  |  |  |  |  |  |  |  |  |
| Lowest | 3,298 | 3,009 | 21.1 | (20-22.3) | 166 | 21.5 | (16.8-26.1) | 26 | 28 | (14.5-41.4) | 97 | 21.7 | (15.5-27.9) |
| Second | 3,394 | 3,122 | 26.1 | (24.8-27.4) | 153 | 23.8 | (17.6-29.9) | 22 | 21.3 | (10-32.7) | 97 | 24.5 | (18-31.1) |
| Third | 3,304 | 3,020 | 26.3 | (25-27.6) | 174 | 27.3 | (21.1-33.5) | 40 | 24.8 | (14.6-35) | 70 | 21.3 | (14.6-28) |
| Highest | 3,326 | 2,987 | 26.5 | (25.2-27.8) | 197 | 27.5 | (22.3-32.6) | 40 | 25.9 | (15.4-36.4) | 102 | 32.5 | (24.3-40.6) |
| Residence urbanization level 4Q |  |  |  |  |  |  |  |  |  |  |  |  |  |
| Lowest (live in rural area) | 3,774 | 3,456 | 31.7 | (30.3-33.1) | 187 | 28.9 | (22.5-35.3) | 38 | 31.3 | (18.7-44) | 93 | 28.1 | (20.6-35.6) |
| Second | 3,285 | 2,954 | 19.1 | (18.1-20.2) | 184 | 21.3 | (16.8-25.7) | 42 | 30 | (17.8-42.1) | 105 | 25.5 | (18.9-32.2) |
| Third | 3,310 | 3,014 | 26.3 | (25.1-27.6) | 185 | 26.8 | (21.6-31.9) | 22 | 13.4 | (6.4-20.3) | 89 | 26.2 | (18.3-34.1) |
| Highest | 3,293 | 3,012 | 22.8 | (21.7-24) | 164 | 23.1 | (17.1-29.1) | 30 | 25.3 | (13.3-37.4) | 87 | 20.1 | (14.4-25.9) |
| Female | 14,376 | 13032 |  |  | 574 |  |  | 222 |  |  | 548 |  |  |
| Type of residence |  |  |  |  |  |  |  |  |  |  |  |  |  |
| Owning house |  |  | 74.4 | (73.1-75.6) |  | 66.6 | (59.4-73.8) |  | 73.6 | (64.5-82.7) |  | 72 | (66.4-77.7) |
| Detached house | 7,390 | 6,741 | 63.8 | (62.4-65.2) | 271 | 53.9 | (46.4-61.5) | 106 | 69.5 | (59.6-79.5) | 272 | 60 | (53.3-66.6) |
| Condominiums | 2,254 | 2,056 | 10.6 | (9.8-11.4) | 94 | 12.2 | (8.7-15.7) | 26 | 6.4 | (1.5-11.3) | 78 | 11.8 | (7.4-16.2) |
| Not owning house |  |  | 25.6 | (24.4-26.9) |  | 33.4 | (26.2-40.6) |  | 26.4 | (17.3-35.5) |  | 28 | (22.3-33.6) |
| Rental housing (Private Apartments) | 3,523 | 3,170 | 18.5 | (17.4-19.5) | 142 | 20.3 | (14.2-26.4) | 70 | 18.3 | (11.3-25.2) | 141 | 18.8 | (14.1-23.4) |
| Rental housing (Public Apartments) | 602 | 534 | 3.8 | (3.3-4.4) | 29 | 6.6 | (2.2-10.9) | 6 | 0.8 | (0-1.6) | 33 | 5.9 | (2.6-9.2) |
| Boarding | 22 | 15 | 0.1 | (0-0.1) | 5 | 0.9 | (-0.3-2.1) | 1 | 0.5 | (-0.5-1.4) | 1 | 0.1 | (-0.1-0.3) |
| Company housing | 324 | 292 | 1.4 | (1.1-1.6) | 19 | 2.8 | (1-4.6) | 7 | 1.8 | (0.1-3.4) | 6 | 1.5 | (0-2.9) |
| Freeload | 17 | 10 | 0.2 | (0-0.4) | 3 | 0.2 | (-0.1-0.5) | 1 | 0.5 | (-0.5-1.6) | 3 | 0.5 | (-0.2-1.1) |
| Temporary house (ie internet cafe) | 1 | 0 |  |  | 1 | 0.1 | (-0.1-0.3) | 0 |  |  | 0 |  |  |
| Others | 243 | 214 | 1.7 | (1.3-2.1) | 10 | 3.1 | (-0.8-7) | 5 | 2.2 | (-0.7-5.2) | 14 | 1.6 | (0.4-2.8) |
| Areal deprivation index (Residence) 4Q |  |  |  |  |  |  |  |  |  |  |  |  |  |
| Lowest | 3,553 | 3,222 | 17.8 | (16.7-18.9) | 152 | 17 | (13-21) | 53 | 15.7 | (6.6-24.8) | 126 | 15 | (10.6-19.4) |
| Second | 3,452 | 3,129 | 24.2 | (22.9-25.5) | 133 | 24.4 | (18.1-30.7) | 60 | 21.9 | (12.6-31.2) | 130 | 21.3 | (15.8-26.9) |
| Third | 3,544 | 3,260 | 29.2 | (27.8-30.6) | 108 | 23.2 | (16.6-29.7) | 52 | 43.3 | (28.9-57.7) | 124 | 24 | (18.1-29.9) |
| Highest | 3,522 | 3,161 | 28.9 | (27.4-30.3) | 152 | 35.5 | (27.7-43.3) | 52 | 19.1 | (11.4-26.8) | 157 | 39.6 | (32.4-46.9) |
| Residence urbanization level 4Q |  |  |  |  |  |  |  |  |  |  |  |  |  |
| Lowest (live in rural area) | 3,849 | 3,519 | 38.3 | (36.8-39.9) | 140 | 35 | (27.3-42.8) | 51 | 26.7 | (16-37.4) | 139 | 40.7 | (33.4-48) |
| Second | 3,520 | 3,169 | 11.5 | (10.7-12.2) | 159 | 13.9 | (10.6-17.2) | 56 | 10.6 | (5.7-15.4) | 136 | 10.6 | (7.7-13.4) |
| Third | 3,543 | 3,214 | 32.7 | (31.2-34.2) | 131 | 34.5 | (26.9-42.1) | 61 | 45.9 | (31.8-60.1) | 137 | 30.6 | (24.1-37.1) |
| Highest | 3,464 | 3,130 | 17.5 | (16.5-18.5) | 144 | 16.6 | (12.4-20.8) | 54 | 16.8 | (8.9-24.6) | 136 | 18.1 | (13.2-23.1) |

Abbreviation: SOGI, sexual orientation and gender identity; CI, confidential interval All %'s represents weighted percentages.

**Table 10.**
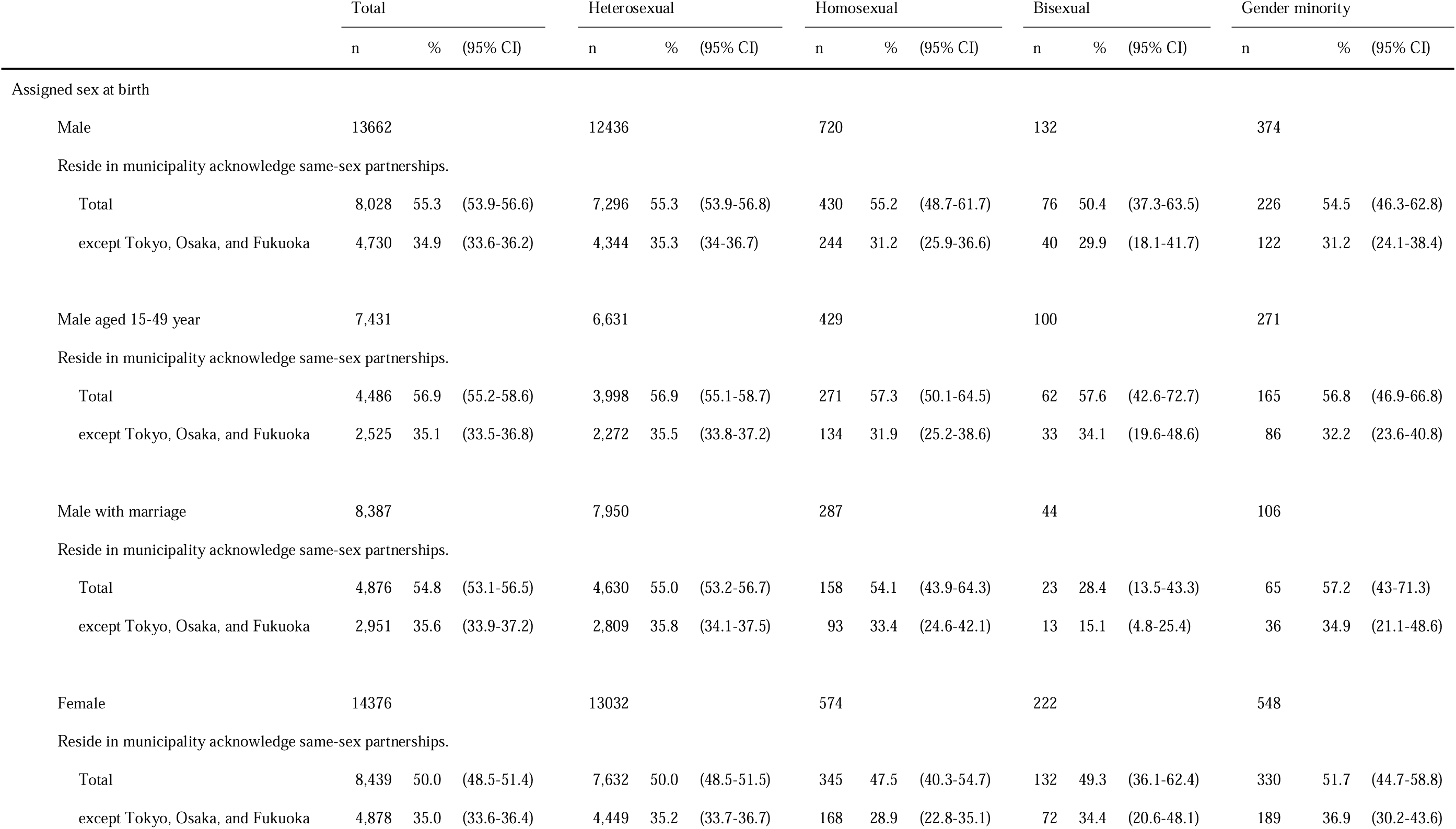
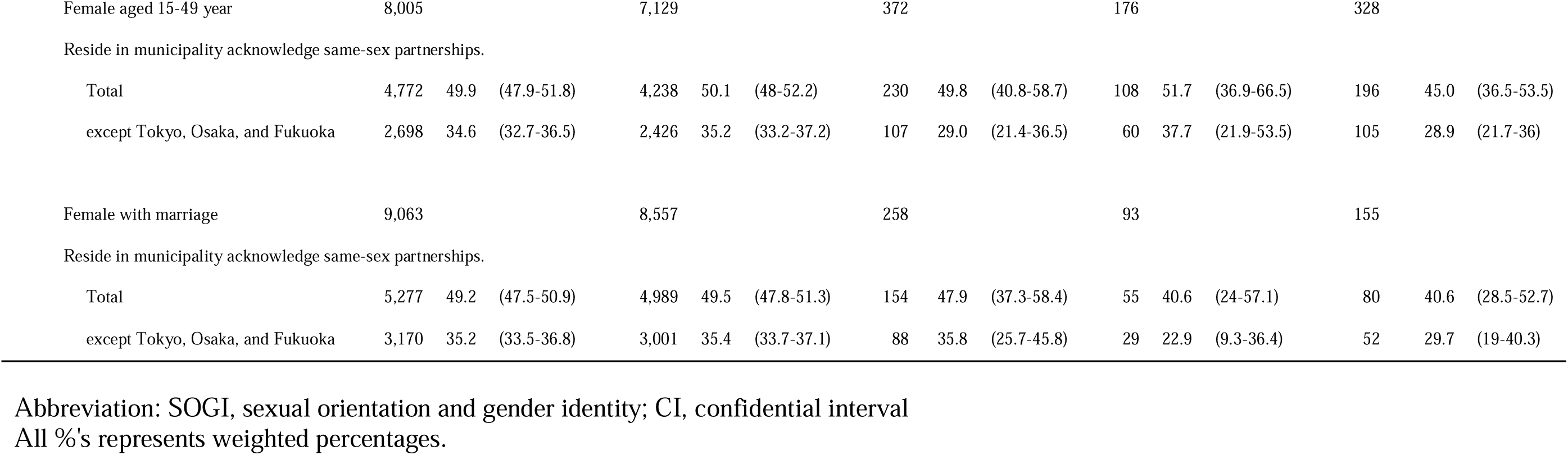
Residents in municipalities that acknowledged same-sex partnerships by SOGI category and sex.

|  | Total |  |  | Heterosexual |  |  | Homosexual |  |  | Bisexual |  |  | Gender minority |  |  |
| --- | --- | --- | --- | --- | --- | --- | --- | --- | --- | --- | --- | --- | --- | --- | --- |
|  | n | % | (95% CI) | n | % | (95% CI) | n | % | (95% CI) | n | % | (95% CI) | n | % | (95% CI) |
| Assigned sex at birth |  |  |  |  |  |  |  |  |  |  |  |  |  |  |  |
| Male | 13662 |  |  | 12436 |  |  | 720 |  |  | 132 |  |  | 374 |  |  |
| Reside in municipality acknowledge same-sex partnerships. |  |  |  |  |  |  |  |  |  |  |  |  |  |  |  |
| Total | 8,028 | 55.3 | (53.9-56.6) | 7,296 | 55.3 | (53.9-56.8) | 430 | 55.2 | (48.7-61.7) | 76 | 50.4 | (37.3-63.5) | 226 | 54.5 | (46.3-62.8) |
| except Tokyo, Osaka, and Fukuoka | 4,730 | 34.9 | (33.6-36.2) | 4,344 | 35.3 | (34-36.7) | 244 | 31.2 | (25.9-36.6) | 40 | 29.9 | (18.1-41.7) | 122 | 31.2 | (24.1-38.4) |
| Male aged 15-49 year |  |  |  |  |  |  |  |  |  |  |  |  |  |  |  |
| Male aged 15-49 year | 7,431 |  |  | 6,631 |  |  | 429 |  |  | 100 |  |  | 271 |  |  |
| Reside in municipality acknowledge same-sex partnerships. |  |  |  |  |  |  |  |  |  |  |  |  |  |  |  |
| Total | 4,486 | 56.9 | (55.2-58.6) | 3,998 | 56.9 | (55.1-58.7) | 271 | 57.3 | (50.1-64.5) | 62 | 57.6 | (42.6-72.7) | 165 | 56.8 | (46.9-66.8) |
| except Tokyo, Osaka, and Fukuoka | 2,525 | 35.1 | (33.5-36.8) | 2,272 | 35.5 | (33.8-37.2) | 134 | 31.9 | (25.2-38.6) | 33 | 34.1 | (19.6-48.6) | 86 | 32.2 | (23.6-40.8) |
| Male with marriage |  |  |  |  |  |  |  |  |  |  |  |  |  |  |  |
| Male with marriage | 8,387 |  |  | 7,950 |  |  | 287 |  |  | 44 |  |  | 106 |  |  |
| Reside in municipality acknowledge same-sex partnerships. |  |  |  |  |  |  |  |  |  |  |  |  |  |  |  |
| Total | 4,876 | 54.8 | (53.1-56.5) | 4,630 | 55.0 | (53.2-56.7) | 158 | 54.1 | (43.9-64.3) | 23 | 28.4 | (13.5-43.3) | 65 | 57.2 | (43-71.3) |
| except Tokyo, Osaka, and Fukuoka | 2,951 | 35.6 | (33.9-37.2) | 2,809 | 35.8 | (34.1-37.5) | 93 | 33.4 | (24.6-42.1) | 13 | 15.1 | (4.8-25.4) | 36 | 34.9 | (21.1-48.6) |
| Female |  |  |  |  |  |  |  |  |  |  |  |  |  |  |  |
| Female | 14376 |  |  | 13032 |  |  | 574 |  |  | 222 |  |  | 548 |  |  |
| Reside in municipality acknowledge same-sex partnerships. |  |  |  |  |  |  |  |  |  |  |  |  |  |  |  |
| Total | 8,439 | 50.0 | (48.5-51.4) | 7,632 | 50.0 | (48.5-51.5) | 345 | 47.5 | (40.3-54.7) | 132 | 49.3 | (36.1-62.4) | 330 | 51.7 | (44.7-58.8) |
| except Tokyo, Osaka, and Fukuoka | 4,878 | 35.0 | (33.6-36.4) | 4,449 | 35.2 | (33.7-36.7) | 168 | 28.9 | (22.8-35.1) | 72 | 34.4 | (20.6-48.1) | 189 | 36.9 | (30.2-43.6) |
| Female aged 15-49 year | 8,005 |  |  | 7,129 |  |  | 372 |  |  | 176 |  |  | 328 |  |  |
| Reside in municipality acknowledge same-sex partnerships. |  |  |  |  |  |  |  |  |  |  |  |  |  |  |  |
| Total | 4,772 | 49.9 | (47.9-51.8) | 4,238 | 50.1 | (48-52.2) | 230 | 49.8 | (40.8-58.7) | 108 | 51.7 | (36.9-66.5) | 196 | 45.0 | (36.5-53.5) |
| except Tokyo, Osaka, and Fukuoka | 2,698 | 34.6 | (32.7-36.5) | 2,426 | 35.2 | (33.2-37.2) | 107 | 29.0 | (21.4-36.5) | 60 | 37.7 | (21.9-53.5) | 105 | 28.9 | (21.7-36) |
| Female with marriage | 9,063 |  |  | 8,557 |  |  | 258 |  |  | 93 |  |  | 155 |  |  |
| Reside in municipality acknowledge same-sex partnerships. |  |  |  |  |  |  |  |  |  |  |  |  |  |  |  |
| Total | 5,277 | 49.2 | (47.5-50.9) | 4,989 | 49.5 | (47.8-51.3) | 154 | 47.9 | (37.3-58.4) | 55 | 40.6 | (24-57.1) | 80 | 40.6 | (28.5-52.7) |
| except Tokyo, Osaka, and Fukuoka | 3,170 | 35.2 | (33.5-36.8) | 3,001 | 35.4 | (33.7-37.1) | 88 | 35.8 | (25.7-45.8) | 29 | 22.9 | (9.3-36.4) | 52 | 29.7 | (19-40.3) |

Abbreviation: SOGI, sexual orientation and gender identity; CI, confidential interval All %'s represents weighted percentages.

**Table 11.** Body mass index by SOGI category and sex.

|  | Total | Heterosexual |  |  | Homosexual |  |  | Bisexual |  |  | Gender minority |  |  |
| --- | --- | --- | --- | --- | --- | --- | --- | --- | --- | --- | --- | --- | --- |
|  | n | n | % | (95% CI) | n | % | (95% CI) | n | % | (95% CI) | n | % | (95% CI) |
| Assigned sex at birth |  |  |  |  |  |  |  |  |  |  |  |  |  |
| Male | 13,662 | 12,436 |  |  | 720 |  |  | 132 |  |  | 374 |  |  |
| 18.5-25 Normal weight | 9,437 | 8,618 | 68.4 | (67.1-69.7) | 490 | 67.9 | (62.5-73.3) | 73 | 54.5 | (41.4-67.7) | 256 | 64.3 | (56.5-72.2) |
| <18.5 Underweight | 788 | 675 | 5.3 | (4.7-6) | 61 | 7.5 | (4.8-10.2) | 11 | 12.9 | (2.7-23.1) | 41 | 13.1 | (7.6-18.7) |
| 25-30 Overweight | 2,841 | 2,617 | 21.6 | (20.4-22.8) | 134 | 16.6 | (12.7-20.5) | 31 | 17.9 | (9.4-26.4) | 59 | 17.3 | (10.9-23.6) |
| >30 Obesity | 596 | 526 | 4.7 | (4.1-5.3) | 35 | 8 | (4.8-11.2) | 17 | 14.7 | (4.1-25.3) | 18 | 5.3 | (1.8-8.7) |
| Female | 14,376 | 13032 |  |  | 574 |  |  | 222 |  |  | 548 |  |  |
| 18.5-25 Normal weight | 9,900 | 9,030 | 68.2 | (66.8-69.6) | 387 | 63 | (55.7-70.4) | 144 | 61.9 | (48.3-75.6) | 339 | 58.4 | (51.3-65.4) |
| <18.5 Underweight | 2,917 | 2,590 | 19 | (17.8-20.1) | 147 | 25.2 | (18.9-31.4) | 51 | 29.3 | (15.3-43.3) | 129 | 26.7 | (20.2-33.3) |
| 25-30 Overweight | 1,242 | 1,138 | 9.9 | (8.9-10.8) | 30 | 10.6 | (4.8-16.5) | 19 | 5.5 | (0.9-10.2) | 55 | 9.9 | (5.7-14.1) |
| >30 Obesity | 317 | 274 | 3 | (2.4-3.5) | 10 | 1.2 | (0.1-2.2) | 8 | 3.2 | (0-6.4) | 25 | 5 | (1.7-8.2) |

Abbreviation: SOGI, sexual orientation and gender identity; CI, confidential interval
All %'s represents weighted percentages.

**Table 12.**
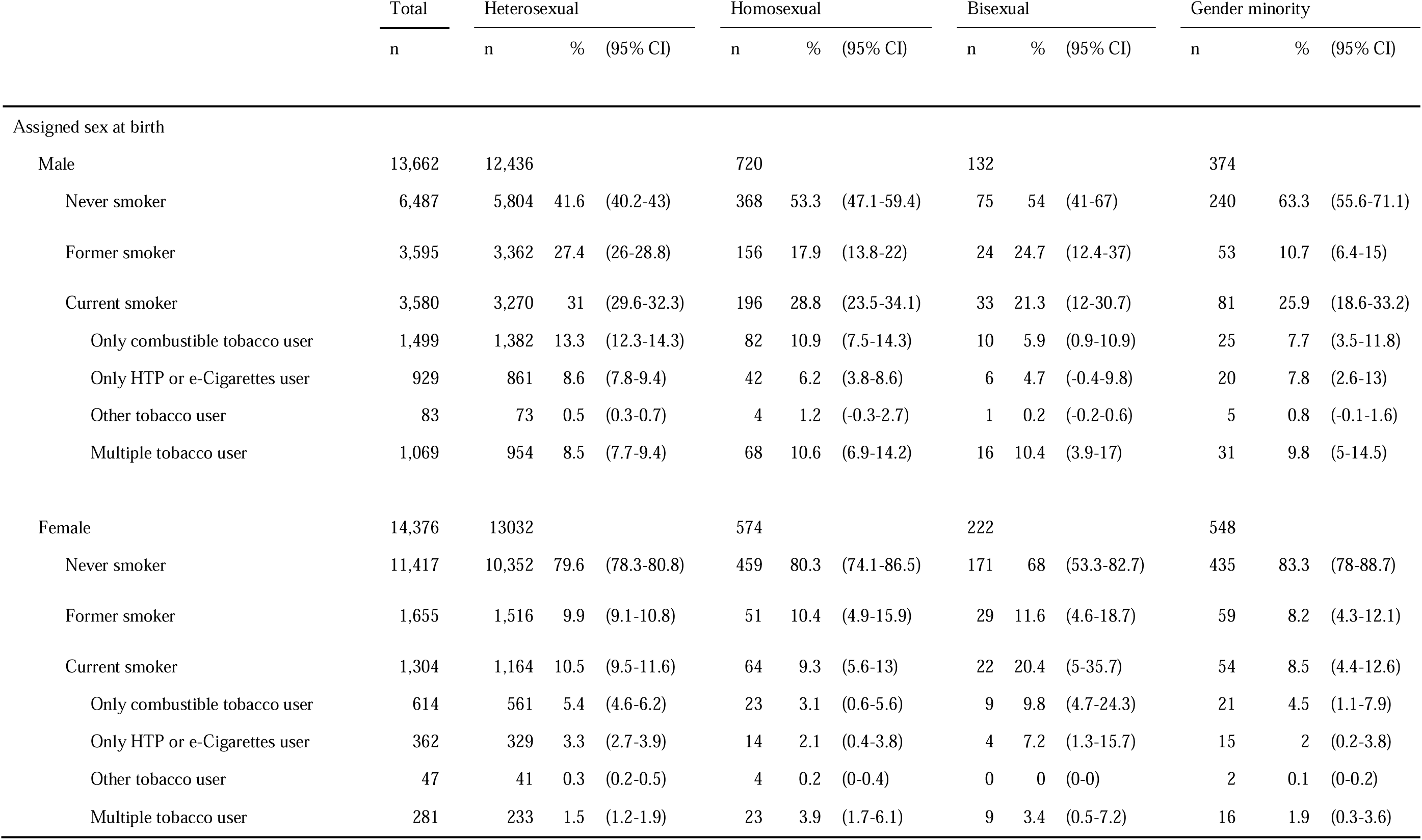
Smoking status by SOGI category and sex.

|  | Total | Heterosexual |  |  | Homosexual |  |  | Bisexual |  |  | Gender minority |  |  |
| --- | --- | --- | --- | --- | --- | --- | --- | --- | --- | --- | --- | --- | --- |
|  | n | n | % | (95% CI) | n | % | (95% CI) | n | % | (95% CI) | n | % | (95% CI) |
| Assigned sex at birth |  |  |  |  |  |  |  |  |  |  |  |  |  |
| Male | 13,662 | 12,436 |  |  | 720 |  |  | 132 |  |  | 374 |  |  |
| Never smoker | 6,487 | 5,804 | 41.6 | (40.2-43) | 368 | 53.3 | (47.1-59.4) | 75 | 54 | (41-67) | 240 | 63.3 | (55.6-71.1) |
| Former smoker | 3,595 | 3,362 | 27.4 | (26-28.8) | 156 | 17.9 | (13.8-22) | 24 | 24.7 | (12.4-37) | 53 | 10.7 | (6.4-15) |
| Current smoker | 3,580 | 3,270 | 31 | (29.6-32.3) | 196 | 28.8 | (23.5-34.1) | 33 | 21.3 | (12-30.7) | 81 | 25.9 | (18.6-33.2) |
| Only combustible tobacco user | 1,499 | 1,382 | 13.3 | (12.3-14.3) | 82 | 10.9 | (7.5-14.3) | 10 | 5.9 | (0.9-10.9) | 25 | 7.7 | (3.5-11.8) |
| Only HTP or e-Cigarettes user | 929 | 861 | 8.6 | (7.8-9.4) | 42 | 6.2 | (3.8-8.6) | 6 | 4.7 | (-0.4-9.8) | 20 | 7.8 | (2.6-13) |
| Other tobacco user | 83 | 73 | 0.5 | (0.3-0.7) | 4 | 1.2 | (-0.3-2.7) | 1 | 0.2 | (-0.2-0.6) | 5 | 0.8 | (-0.1-1.6) |
| Multiple tobacco user | 1,069 | 954 | 8.5 | (7.7-9.4) | 68 | 10.6 | (6.9-14.2) | 16 | 10.4 | (3.9-17) | 31 | 9.8 | (5-14.5) |
| Female | 14,376 | 13,032 |  |  | 574 |  |  | 222 |  |  | 548 |  |  |
| Never smoker | 11,417 | 10,352 | 79.6 | (78.3-80.8) | 459 | 80.3 | (74.1-86.5) | 171 | 68 | (53.3-82.7) | 435 | 83.3 | (78-88.7) |
| Former smoker | 1,655 | 1,516 | 9.9 | (9.1-10.8) | 51 | 10.4 | (4.9-15.9) | 29 | 11.6 | (4.6-18.7) | 59 | 8.2 | (4.3-12.1) |
| Current smoker | 1,304 | 1,164 | 10.5 | (9.5-11.6) | 64 | 9.3 | (5.6-13) | 22 | 20.4 | (5-35.7) | 54 | 8.5 | (4.4-12.6) |
| Only combustible tobacco user | 614 | 561 | 5.4 | (4.6-6.2) | 23 | 3.1 | (0.6-5.6) | 9 | 9.8 | (4.7-24.3) | 21 | 4.5 | (1.1-7.9) |
| Only HTP or e-Cigarettes user | 362 | 329 | 3.3 | (2.7-3.9) | 14 | 2.1 | (0.4-3.8) | 4 | 7.2 | (1.3-15.7) | 15 | 2 | (0.2-3.8) |
| Other tobacco user | 47 | 41 | 0.3 | (0.2-0.5) | 4 | 0.2 | (0-0.4) | 0 | 0 | (0-0) | 2 | 0.1 | (0-0.2) |
| Multiple tobacco user | 281 | 233 | 1.5 | (1.2-1.9) | 23 | 3.9 | (1.7-6.1) | 9 | 3.4 | (0.5-7.2) | 16 | 1.9 | (0.3-3.6) |

**Table 13.** Substance use by SOGI category and sex.

|  | Total | Heterosexual |  |  | Homosexual |  |  | Bisexual |  |  | Gender minority |  |  |
| --- | --- | --- | --- | --- | --- | --- | --- | --- | --- | --- | --- | --- | --- |
|  | n | n | % | (95% CI) | n | % | (95% CI) | n | % | (95% CI) | n | % | (95% CI) |
| Assigned sex at birth |  |  |  |  |  |  |  |  |  |  |  |  |  |
| Male | 13,662 | 12,436 |  |  | 720 |  |  | 132 |  |  | 374 |  |  |
| Alcohol consumption |  |  |  |  |  |  |  |  |  |  |  |  |  |
| Never drinker | 4,292 | 3793 | 40.9 | (39.4-42.4) | 276 | 49.9 | (43.6-56.2) | 50 | 53.9 | (41.3-66.6) | 173 | 57.7 | (50-65.4) |
| Former drinker | 1,073 | 937 | 7.4 | (6.6-8.2) | 81 | 9.5 | (6.3-12.8) | 16 | 10.9 | (4-17.7) | 39 | 7.9 | (4.1-11.7) |
| Habitual drinker | 8,297 | 7706 | 51.7 | (50.2-53.1) | 363 | 40.6 | (34.8-46.4) | 66 | 35.2 | (23.8-46.5) | 162 | 34.4 | (27.2-41.5) |
| Sedatives or tranquilizers |  |  |  |  |  |  |  |  |  |  |  |  |  |
| Never user | 11,958 | 10,948 | 88.5 | (87.5-89.4) | 597 | 85.2 | (81.4-89) | 103 | 84.1 | (76.8-91.3) | 310 | 84.4 | (78.6-90.2) |
| Former user | 557 | 480 | 3.5 | (3-3.9) | 46 | 5.6 | (3.2-8) | 10 | 6.7 | (1.8-11.6) | 21 | 5.9 | (1.4-10.5) |
| Current user | 1,147 | 1,008 | 8.1 | (7.2-8.9) | 77 | 9.2 | (6.1-12.2) | 19 | 9.2 | (3.9-14.5) | 43 | 9.7 | (5.6-13.7) |
| Non-prescription opioids |  |  |  |  |  |  |  |  |  |  |  |  |  |
| Never user | 13,425 | 12,279 | 98.7 | (98.4-99) | 666 | 92.2 | (89.1-95.4) | 125 | 94.2 | (88.3-100) | 355 | 93.8 | (89.1-98.4) |
| Former user | 139 | 92 | 0.8 | (0.6-1.1) | 30 | 5.2 | (2.3-8.2) | 5 | 3.8 | (-0.8-8.4) | 12 | 5.4 | (0.8-10) |
| Current user | 98 | 65 | 0.4 | (0.3-0.6) | 24 | 2.5 | (1.2-3.9) | 2 | 2.1 | (-1.7-5.9) | 7 | 0.8 | (0.1-1.6) |
| Organic solvents |  |  |  |  |  |  |  |  |  |  |  |  |  |
| Never user | 13,396 | 12,261 | 98.5 | (98.2-98.9) | 659 | 91.8 | (88.8-94.9) | 122 | 94.1 | (89-99.2) | 354 | 91.1 | (85.6-96.6) |
| Former user | 176 | 120 | 1.1 | (0.8-1.3) | 37 | 6.0 | (3.2-8.8) | 7 | 5.0 | (0.1-10) | 12 | 4.0 | (0-8.1) |
| Current user | 90 | 55 | 0.4 | (0.3-0.6) | 24 | 2.2 | (1-3.4) | 3 | 0.9 | (-0.3-2) | 8 | 4.9 | (0.8-9) |
| Illicit drug |  |  |  |  |  |  |  |  |  |  |  |  |  |
| Never user | 13,427 | 12,282 | 98.5 | (98.2-98.8) | 667 | 93.4 | (90.6-96.2) | 124 | 92.6 | (86.3-99) | 354 | 93.7 | (89.9-97.6) |
| Former user | 153 | 102 | 1.0 | (0.7-1.3) | 35 | 5.0 | (2.4-7.6) | 5 | 3.3 | (-0.9-7.5) | 11 | 4.1 | (0.7-7.4) |
| Current user | 82 | 52 | 0.5 | (0.3-0.6) | 18 | 1.6 | (0.6-2.5) | 3 | 4.1 | (-0.8-8.9) | 9 | 2.2 | (0.2-4.3) |
| Cannabis (Marijuana) |  |  |  |  |  |  |  |  |  |  |  |  |  |
| Never user | 13,404 | 12,258 | 98.5 | (98.2-98.8) | 671 | 94.3 | (91.9-96.7) | 123 | 92.5 | (86.3-98.7) | 352 | 90.6 | (85-96.1) |
| Former user | 155 | 107 | 1.0 | (0.7-1.2) | 28 | 2.8 | (1.1-4.5) | 7 | 4.9 | (0.2-9.7) | 13 | 4.4 | (0.8-7.9) |
| Current user | 103 | 71 | 0.6 | (0.4-0.8) | 21 | 2.9 | (1.2-4.7) | 2 | 2.6 | (-1.6-6.8) | 9 | 5.1 | (0.5-9.6) |
| Stimulants/Cocaine/Heroin |  |  |  |  |  |  |  |  |  |  |  |  |  |
| Never user | 13,443 | 12,293 | 98.7 | (98.3-99) | 669 | 94.7 | (92.8-96.6) | 124 | 93.0 | (86.8-99.1) | 357 | 93.0 | (88-98) |
| Former user | 138 | 90 | 0.9 | (0.6-1.2) | 33 | 3.0 | (1.7-4.3) | 5 | 5.5 | (-0.3-11.3) | 10 | 1.8 | (0.3-3.2) |
| Current user | 81 | 53 | 0.4 | (0.3-0.6) | 18 | 2.3 | (0.9-3.7) | 3 | 1.6 | (-0.5-3.7) | 7 | 5.3 | (0.4-10.1) |
| Female | 14,376 | 13,032 |  |  | 574 |  |  | 222 |  |  | 548 |  |  |
| Alcohol consumption |  |  |  |  |  |  |  |  |  |  |  |  |  |
| Never drinker | 7,055 | 6,317 | 69.0 | (67.8-70.2) | 298 | 72.4 | (66.9-77.8) | 114 | 76.4 | (67.8-85) | 326 | 73.6 | (68-79.2) |
| Former drinker | 1,336 | 1,229 | 5.7 | (5.2-6.2) | 50 | 6.3 | (3.3-9.4) | 20 | 4.0 | (1.5-6.6) | 37 | 3.4 | (1.7-5) |
| Habitual drinker | 5,985 | 5,486 | 25.3 | (24.2-26.4) | 226 | 21.3 | (16.7-25.9) | 88 | 19.6 | (11.6-27.5) | 185 | 23.1 | (17.7-28.4) |
| Sedatives or tranquilizers |  |  |  |  |  |  |  |  |  |  |  |  |  |
| Never user | 12426 | 11341 | 87.1 | (86.1-88.1) | 485 | 82.7 | (77.3-88.1) | 165 | 81.2 | (73.5-88.9) | 435 | 80.4 | (74.9-86) |
| Former user | 654 | 569 | 4.2 | (3.7-4.8) | 32 | 8.7 | (4.1-13.4) | 23 | 6.4 | (2.4-10.5) | 30 | 5.5 | (2.3-8.6) |
| Current user | 1296 | 1122 | 8.6 | (7.8-9.5) | 57 | 8.6 | (5.3-11.8) | 34 | 12.4 | (6-18.8) | 83 | 14.1 | (9.2-19) |
| Non-prescription opioids |  |  |  |  |  |  |  |  |  |  |  |  |  |
| Never user | 14262 | 12958 | 99.4 | (99.1-99.7) | 548 | 95.3 | (92.7-97.8) | 213 | 95.9 | (92.2-99.6) | 543 | 99.8 | (99.7-100) |
| Former user | 66 | 43 | 0.4 | (0.2-0.7) | 17 | 4.0 | (1.5-6.4) | 5 | 2.4 | (-0.5-5.3) | 1 | 0.0 | (0-0.1) |
| Current user | 48 | 31 | 0.2 | (0.1-0.3) | 9 | 0.8 | (0.2-1.4) | 4 | 1.7 | (-0.6-4) | 4 | 0.2 | (0-0.3) |
| Organic solvents |  |  |  |  |  |  |  |  |  |  |  |  |  |
| Never user | 14251 | 12954 | 99.3 | (99-99.6) | 548 | 95.4 | (92.9-97.9) | 213 | 96.1 | (92.5-99.8) | 536 | 98.0 | (95.6-100.5) |
| Former user | 75 | 50 | 0.4 | (0.2-0.6) | 15 | 3.2 | (1-5.4) | 6 | 2.6 | (-0.3-5.5) | 4 | 0.3 | (0-0.6) |
| Current user | 50 | 28 | 0.3 | (0-0.5) | 11 | 1.5 | (0.3-2.6) | 3 | 1.3 | (-0.9-3.5) | 8 | 1.7 | (-0.8-4.1) |
| Illicit drug |  |  |  |  |  |  |  |  |  |  |  |  |  |
| Never user | 14257 | 12957 | 99.5 | (99.2-99.7) | 547 | 95.4 | (93-97.8) | 214 | 97.5 | (94.7-100.3) | 539 | 99.5 | (99.1-99.9) |
| Former user | 66 | 41 | 0.3 | (0.1-0.5) | 16 | 2.9 | (1-4.9) | 4 | 1.7 | (-1-4.3) | 5 | 0.4 | (0-0.7) |
| Current user | 53 | 34 | 0.2 | (0.1-0.3) | 11 | 1.6 | (0.2-3.1) | 4 | 0.9 | (-0.1-1.9) | 4 | 0.1 | (0-0.2) |
| Cannabis (Marijuana) |  |  |  |  |  |  |  |  |  |  |  |  |  |
| Never user | 14258 | 12956 | 99.6 | (99.4-99.7) | 548 | 96.4 | (94.4-98.4) | 216 | 97.1 | (93.7-100.5) | 538 | 99.6 | (99.4-99.9) |
| Former user | 69 | 46 | 0.3 | (0.2-0.5) | 13 | 1.4 | (0.4-2.3) | 4 | 2.7 | (-0.7-6.1) | 6 | 0.2 | (0-0.4) |
| Current user | 49 | 30 | 0.1 | (0.1-0.2) | 13 | 2.2 | (0.4-4) | 2 | 0.1 | (-0.1-0.3) | 4 | 0.2 | (0-0.3) |
| Stimulants/Cocaine/Heroin |  |  |  |  |  |  |  |  |  |  |  |  |  |
| Never user | 14269 | 12966 | 99.4 | (99.1-99.7) | 549 | 95.9 | (93.7-98.2) | 216 | 97.8 | (95.1-100.6) | 538 | 99.6 | (99.2-99.9) |
| Former user | 60 | 36 | 0.4 | (0.1-0.7) | 15 | 2.2 | (0.5-3.8) | 5 | 0.8 | (-0.1-1.7) | 4 | 0.2 | (0-0.4) |
| Current user | 47 | 30 | 0.2 | (0.1-0.3) | 10 | 1.9 | (0.4-3.4) | 1 | 1.3 | (-1.3-3.9) | 6 | 0.3 | (0-0.5) |

Abbreviation: SOGI, sexual orientation and gender identity; CI, confidential interval All %'s represents weighted percentages.

**Table 14.** Comorbidity by SOGI category and sex.

|  | Total | Heterosexual |  |  | Gay/Lesbian |  |  | Bisexual |  |  | Gender minority |  |  |
| --- | --- | --- | --- | --- | --- | --- | --- | --- | --- | --- | --- | --- | --- |
|  | n | n | % | (95% CI) | n | % | (95% CI) | n | % | (95% CI) | n | % | (95% CI) |
| Assigned sex at birth |  |  |  |  |  |  |  |  |  |  |  |  |  |
| Male | 13,662 | 12,436 |  |  | 720 |  |  | 132 |  |  | 374 |  |  |
| Metabolic syndrome |  |  |  |  |  |  |  |  |  |  |  |  |  |
| Hypertension | 3,537 | 3,269 | 26.6 | (25.2-27.9) | 166 | 21.9 | (16.1-27.7) | 34 | 22.8 | (11.4-34.3) | 68 | 18.9 | (12.7-25) |
| Diabetes mellitus | 1,295 | 1,184 | 10.9 | (9.9-12) | 59 | 6.6 | (3.8-9.4) | 17 | 18.5 | (6.7-30.2) | 35 | 9.2 | (4.6-13.8) |
| Dyslipidemia | 2,179 | 2,012 | 16.2 | (15.1-17.3) | 100 | 14.0 | (8.4-19.6) | 19 | 9.7 | (3.5-15.9) | 48 | 13.1 | (7.3-18.9) |
| Pneumonia/bronchitis | 250 | 183 | 1.7 | (1.3-2.2) | 40 | 5.0 | (2.5-7.5) | 8 | 4.0 | (-0.3-8.3) | 19 | 8.8 | (3.2-14.4) |
| Allergic disease |  |  |  |  |  |  |  |  |  |  |  |  |  |
| Asthma | 442 | 363 | 3.0 | (2.5-3.5) | 46 | 6.3 | (3.6-8.9) | 9 | 11.2 | (2.8-19.7) | 24 | 8.6 | (3.4-13.7) |
| Atopic dermatitis | 696 | 591 | 4.4 | (3.8-4.9) | 57 | 7.1 | (4.1-10.2) | 9 | 5.4 | (0.7-10.2) | 39 | 15.1 | (7.5-22.7) |
| Allergic rhinitis | 2,751 | 2,517 | 19.5 | (18.4-20.6) | 120 | 17.1 | (12.6-21.7) | 36 | 21.3 | (12.5-30.2) | 78 | 22.5 | (15.5-29.4) |
| Dental disease |  |  |  |  |  |  |  |  |  |  |  |  |  |
| Periodontal disease | 2,083 | 1,901 | 16.0 | (14.9-17.1) | 110 | 18.5 | (11.7-25.3) | 15 | 11.3 | (3.8-18.9) | 57 | 16.3 | (10.3-22.3) |
| Caries | 1,874 | 1,685 | 14.3 | (13.3-15.3) | 111 | 16.5 | (10.9-22.1) | 20 | 16.7 | (7.8-25.5) | 58 | 17.1 | (10.9-23.4) |
| Cataract | 633 | 547 | 4.0 | (3.5-4.6) | 51 | 5.2 | (3.1-7.3) | 5 | 2.9 | (-1-6.8) | 30 | 9.6 | (4.3-14.8) |
| Cardio-cerebrovascular disease |  |  |  |  |  |  |  |  |  |  |  |  |  |
| Angina pectoris/myocardial infarction | 403 | 331 | 3.2 | (2.7-3.8) | 39 | 10.8 | (3.8-17.8) | 9 | 5.7 | (0.8-10.6) | 24 | 9.0 | (3.6-14.4) |
| Stroke, Cerebral infarction/hemorrhage | 204 | 149 | 1.3 | (1-1.7) | 31 | 5.7 | (0.5-10.8) | 6 | 5.1 | (0.1-10) | 18 | 5.5 | (1.9-9.2) |
| Chronic obstructive pulmonary disease | 191 | 138 | 1.3 | (0.9-1.7) | 28 | 4.0 | (1.8-6.2) | 7 | 4.9 | (0.2-9.5) | 18 | 7.4 | (2.3-12.6) |
| Chronic kidney disease | 289 | 222 | 1.8 | (1.4-2.2) | 33 | 4.8 | (2.3-7.3) | 12 | 10.5 | (3-18) | 22 | 10.2 | (4.3-16.1) |
| Chronic hepatitis/Cirrhosis | 221 | 163 | 1.2 | (0.9-1.5) | 31 | 3.8 | (1.7-5.9) | 10 | 6.4 | (1.3-11.5) | 17 | 6.7 | (1.7-11.6) |
| Immunodeficiency disease | 230 | 181 | 1.7 | (1.3-2.1) | 32 | 2.8 | (1.2-4.4) | 5 | 1.8 | (-0.2-3.8) | 12 | 6.2 | (1.3-11.2) |
| Cancer/Malignancy | 300 | 247 | 2.3 | (1.7-2.8) | 27 | 2.7 | (1.1-4.3) | 10 | 7.6 | (1.7-13.6) | 16 | 6.6 | (1.9-11.4) |
| Chronic pain/headache/backache | 2,175 | 2,005 | 16.7 | (15.6-17.7) | 82 | 9.2 | (6.2-12.1) | 23 | 15.9 | (7.5-24.4) | 65 | 18.1 | (11.8-24.3) |
| Mental disease |  |  |  |  |  |  |  |  |  |  |  |  |  |
| Depression | 647 | 532 | 4.0 | (3.4-4.6) | 54 | 5.5 | (3.3-7.8) | 18 | 7.8 | (3-12.6) | 43 | 11.9 | (6.5-17.3) |
| Psychiatric disorders other than depression | 539 | 434 | 3.8 | (3.1-4.4) | 41 | 4.5 | (2.3-6.7) | 22 | 12.3 | (5.9-18.8) | 42 | 15.1 | (8.9-21.3) |
| Female |  |  |  |  |  |  |  |  |  |  |  |  |  |
| Female | 14,376 | 13032 |  |  | 574 |  |  | 222 |  |  | 548 |  |  |
| Metabolic syndrome |  |  |  |  |  |  |  |  |  |  |  |  |  |
| Hypertension | 2021 | 1840 | 15.8 | (14.7-17) | 81 | 17.8 | (11.2-24.4) | 25 | 9.0 | (3.5-14.5) | 75 | 15.8 | (10.3-21.2) |
| Diabetes mellitus | 487 | 421 | 4.0 | (3.4-4.5) | 28 | 7.7 | (3.1-12.4) | 10 | 3.9 | (0.5-7.2) | 28 | 3.5 | (1.5-5.5) |
| Dyslipidemia | 1875 | 1719 | 13.9 | (12.8-14.9) | 60 | 9.5 | (5.6-13.4) | 28 | 8.8 | (3.4-14.2) | 68 | 11.5 | (7-15.9) |
| Pneumonia/bronchitis | 185 | 148 | 1.4 | (1-1.8) | 18 | 3.9 | (1.6-6.2) | 4 | 1.6 | (-1-4.2) | 15 | 1.8 | (0.5-3.1) |
| Allergic disease |  |  |  |  |  |  |  |  |  |  |  |  |  |
| Asthma | 483 | 422 | 3.6 | (3-4.2) | 27 | 4.4 | (2.2-6.6) | 10 | 1.8 | (0.3-3.2) | 24 | 3.2 | (1.2-5.3) |
| Atopic dermatitis | 807 | 697 | 4.9 | (4.3-5.6) | 49 | 10.2 | (5.5-15) | 13 | 4.1 | (0.7-7.5) | 48 | 8.6 | (5.1-12.1) |
| Allergic rhinitis | 2845 | 2574 | 19.5 | (18.3-20.7) | 100 | 15.2 | (10.2-20.1) | 51 | 27.7 | (13.5-41.8) | 120 | 21.4 | (15.8-27) |
| Dental disease |  |  |  |  |  |  |  |  |  |  |  |  |  |
| Periodontal disease | 2187 | 1993 | 16.4 | (15.3-17.6) | 76 | 15.5 | (9.8-21.3) | 38 | 22.1 | (8-36.2) | 80 | 15.3 | (9.8-20.8) |
| Caries | 1,766 | 1,548 | 12.9 | (11.8-13.9) | 80 | 16.7 | (10.8-22.6) | 45 | 18.7 | (10.4-27) | 93 | 15.5 | (10.2-20.8) |
| Cataract | 700 | 618 | 5.3 | (4.6-6) | 37 | 7.2 | (3.6-10.8) | 5 | 0.9 | (-0.1-1.8) | 40 | 8.2 | (4.1-12.3) |
| Cardio-cerebrovascular disease |  |  |  |  |  |  |  |  |  |  |  |  |  |
| Angina pectoris/myocardial infarction | 160 | 124 | 1.3 | (0.9-1.7) | 20 | 4.7 | (1.4-8) | 5 | 2.0 | (-0.8-4.7) | 11 | 1.2 | (0.2-2.2) |
| Stroke, Cerebral infarction/hemorrhage | 100 | 74 | 0.7 | (0.4-0.9) | 16 | 3.5 | (1.3-5.6) | 2 | 0.9 | (-0.4-2.3) | 8 | 0.4 | (0.1-0.8) |
| Chronic obstructive pulmonary disease | 72 | 47 | 0.3 | (0.2-0.5) | 13 | 2.4 | (0.5-4.2) | 5 | 2.2 | (-0.6-4.9) | 7 | 0.4 | (0.1-0.8) |
| Chronic kidney disease | 168 | 135 | 1.4 | (1-1.8) | 17 | 2.5 | (0.7-4.3) | 4 | 2.1 | (-0.7-4.8) | 12 | 1.1 | (0.2-1.9) |
| Chronic hepatitis/Cirrhosis | 114 | 87 | 0.6 | (0.4-0.8) | 12 | 1.9 | (0.2-3.7) | 4 | 2.0 | (-0.7-4.7) | 11 | 0.6 | (0.2-1) |
| Immunodeficiency disease | 320 | 278 | 2.4 | (2-2.8) | 18 | 4.0 | (0.9-7.1) | 10 | 5.5 | (0.4-10.5) | 14 | 2.1 | (0.5-3.7) |
| Cancer/Malignancy | 280 | 246 | 2.1 | (1.6-2.5) | 15 | 2.1 | (0.4-3.9) | 3 | 1.8 | (-0.9-4.5) | 16 | 2.1 | (-0.1-4.4) |
| Chronic pain/headache/backache | 2,726 | 2,445 | 20.4 | (19.2-21.7) | 83 | 16.3 | (10.9-21.7) | 54 | 21.5 | (12.8-30.3) | 144 | 28.1 | (21.8-34.3) |
| Mental disease |  |  |  |  |  |  |  |  |  |  |  |  |  |
| Depression | 574 | 461 | 3.7 | (3-4.3) | 37 | 5.4 | (3.1-7.8) | 17 | 9.9 | (3.1-16.7) | 59 | 10.1 | (6.3-13.8) |
| Psychiatric disorders other than depression | 631 | 500 | 4.6 | (3.9-5.3) | 40 | 6.0 | (3.3-8.7) | 25 | 8.1 | (3.7-12.5) | 66 | 8.7 | (5.5-11.8) |

Abbreviation: SOGI, sexual orientation and gender identity; CI, confidential interval All %'s represents weighted percentages.

**Table 15.** Health-related and COVID-19-related variables by SOGI category and sex.

|  | Total |  | Heterosexual |  |  | Homosexual |  |  | Bisexual |  |  | Gender minority |  |  |
| --- | --- | --- | --- | --- | --- | --- | --- | --- | --- | --- | --- | --- | --- | --- |
|  | n |  | n | % | (95% CI) | n | % | (95% CI) | n | % | (95% CI) | n | % | (95% CI) |
| Assigned sex at birth |  |  |  |  |  |  |  |  |  |  |  |  |  |  |
| Male | 13,662 |  | 12,436 |  |  | 720 |  |  | 132 |  |  | 374 |  |  |
| Self-rated health, good | 7,592 |  | 7,001 | 41.2 | (39.9-42.5) | 362 | 36.4 | (31-41.7) | 59 | 33.8 | (22.9-44.8) | 170 | 38.9 | (31-46.9) |
| Serious psychological distress | 1,319 |  | 1,096 | 8.2 | (7.4-9.0) | 111 | 16 | (11.8-20.1) | 32 | 16.2 | (8.6-23.9) | 80 | 19.2 | (12.1-26.4) |
| Short-term UCLA Loneliness Scale* | 5.6 | (5.5-5.6) |  | 5.5 | (5.5-5.6) |  | 5.2 | (4.9-5.5) |  | 6.8 | (6.1-7.5) |  | 6.3 | (5.8-6.9) |
| Fear of COVID-19 Scale* | 17.0 | (16.9-17.2) |  | 16.9 | (16.8-17.0) |  | 18.2 | (17.4-19.0) |  | 17 | (16.0-18.1) |  | 18.8 | (17.8-19.8) |
| COVID-19 vaccination status |  |  |  |  |  |  |  |  |  |  |  |  |  |  |
| Not fully vaccinated | 1,567 |  | 1,335 | 12 | (11.1-13) | 135 | 20.7 | (14.9-26.4) | 17 | 18.6 | (5.7-31.4) | 77 | 18.8 | (13.4-24.1) |
| Fully vaccinated | 1,737 |  | 1,559 | 13.1 | (12.1-14) | 98 | 16.4 | (11.9-21) | 25 | 19.9 | (9.8-30) | 55 | 15.3 | (9.5-21) |
| Boosted vaccinated | 10,361 |  | 10,361 | 74.9 | (73.7-76.1) | 487 | 62.9 | (56.6-69.2) | 90 | 61.5 | (48.1-75) | 242 | 66 | (58.7-73.2) |
| Female | 14,376 |  | 13032 |  |  | 574 |  |  | 222 |  |  | 548 |  |  |
| Self-rated health, good | 8,265 |  | 7,620 | 38.8 | (37.4-40.3) | 327 | 38.6 | (31.9-45.2) | 95 | 30 | (18.7-41.3) | 223 | 29.2 | (23.4-35.1) |
| Serious psychological distress | 1,457 |  | 1,195 | 10.4 | (9.3-11.4) | 88 | 15.4 | (10.4-20.4) | 61 | 26.9 | (16.2-37.6) | 113 | 17.9 | (13.1-22.6) |
| Short-term UCLA Loneliness Scale* | 6.0 | (6.0-6.1) |  | 6.0 | (5.9-6.1) |  | 5.8 | (5.4-6.2) |  | 7.2 | (6.5-7.9) |  | 6.8 | (6.4-7.2) |
| Fear of COVID-19 Scale* | 17.8 | (17.7-18.0) |  | 17.8 | (17.6-18.0) |  | 19.4 | (18.4-20.3) |  | 17.6 | (16.4-18.8) |  | 17 | (16.3-17.7) |
| COVID-19 vaccination status |  |  |  |  |  |  |  |  |  |  |  |  |  |  |
| Not fully vaccinated | 1,719 |  | 1,512 | 12.7 | (11.6-13.8) | 89 | 16.7 | (10.5-23) | 40 | 19.1 | (9.3-29) | 78 | 14.6 | (9.6-19.6) |
| Fully vaccinated | 2,089 |  | 1,881 | 14.7 | (13.7-15.8) | 86 | 14.8 | (9.9-19.7) | 45 | 25.5 | (14.9-36) | 77 | 18.3 | (12.3-24.4) |
| Boosted vaccinated | 10,568 |  | 9,639 | 72.5 | (71.1-73.9) | 399 | 68.5 | (61.4-75.6) | 137 | 55.4 | (42.6-68.2) | 393 | 67.1 | (60.2-73.9) |

Abbreviation: SOGI, sexual orientation and gender identity; CI, confidential interval
All %'s represents weighted percentages.

SGMs were less likely to be married or in a partnership compared to heterosexuals (65.1% heterosexuals, 43.3% homosexuals, 39.1% bisexuals, 26.7% GMs). More male same-sex couples existed compared to females (56.3% vs. 33.3%). In bisexual and GM groups, same-sex marriage was higher in males (6.7% and 7.2%) compared to females (0% and 0.7%), with most females in opposite-sex marriages. Same-sex marriage was more common in reproductive-aged SGMs. No generational differences were observed for GMs (**Table 2** & Supplementary Table 2).

SGMs had higher proportions of single households (14.9% heterosexuals, 27.2% homosexuals, 17.1% bisexuals, 28.9% GMs). Bisexuals had fewer parenting households than heterosexuals, homosexuals, and GMs (31.0% vs. 43.9%, 45.1%, and 39.6%).

Married/partnered male SGMs had higher parenting rates across all categories, with no female bisexual same-sex marriages. Married/partnered male GMs and female homosexuals had higher parenting proportions in same-sex than opposite-sex marriages (56.9% vs 30.8% and 42.5% vs 31.0%, respectively) (**Table 2** & Supplementary Table 3).

SGMs had lower household equivalent income than heterosexuals (354 [JPY/10,000] heterosexuals, 322 homosexuals, 312 bisexuals, 294 GMs), even if divided by assigned sex at birth. Female GMs and male bisexuals had the lowest income among the same assigned sexes.

High-income quintiles (4th and 5th quintiles) were least common among GMs, and the lowest-income quintile was more common across all SGMs than heterosexuals (**Table 2**, Supplementary Table 4, **Figure 3**).

**Figure 3.**
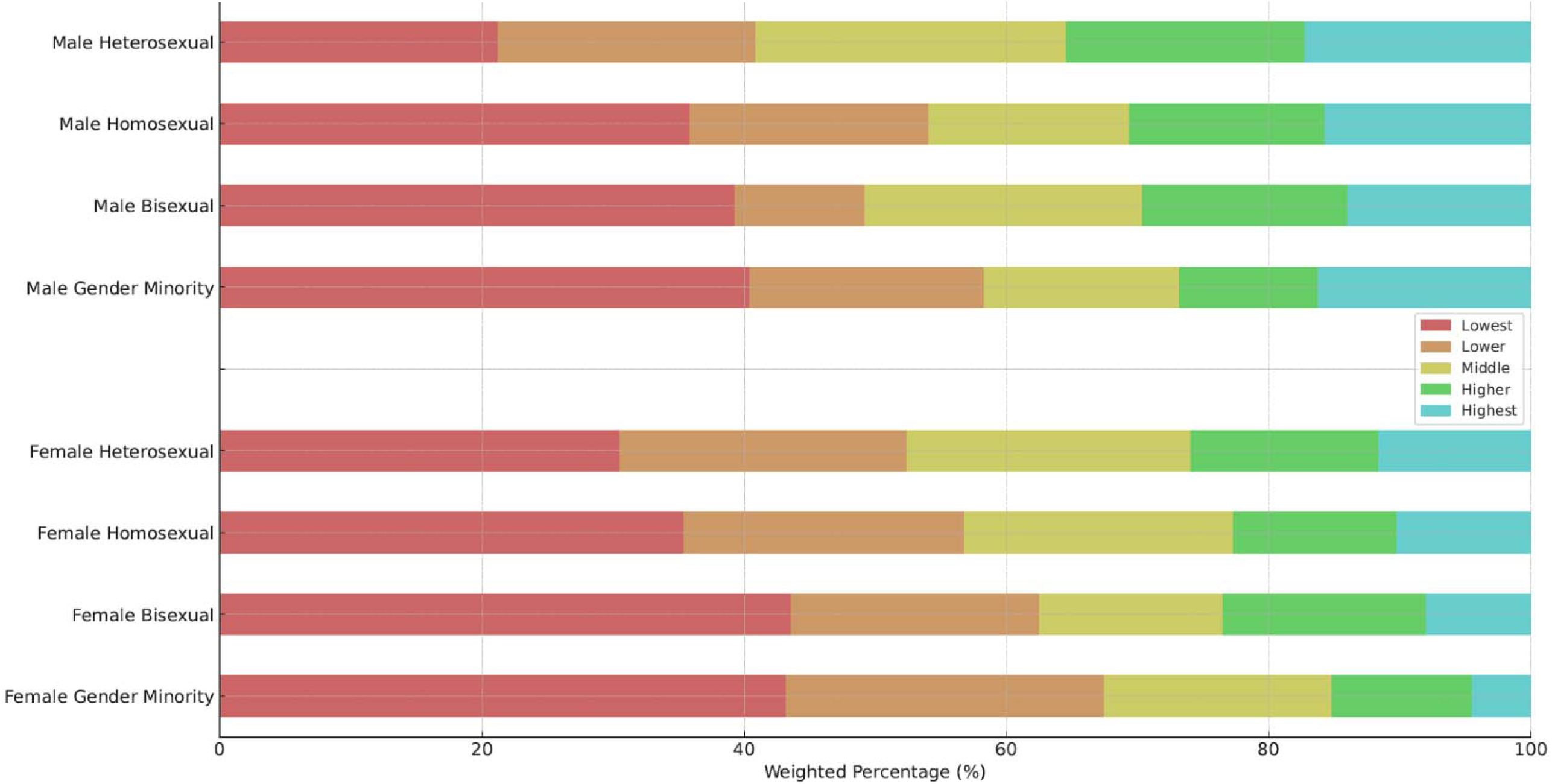
Household equivalent income by SOGI category and sex. Abbreviation: SOGI, sexual orientation and gender identity

Degree holders were less prevalent among SGMs than heterosexuals, with females outnumbering males in each category (46.9% vs. 42.8% heterosexuals, 49.3% vs. 38.1% homosexuals, 39.5% vs. 37.4% bisexuals, 44.2% vs. 35.3% GMs). In any SGM category, bachelor’s and higher degree holders showed little difference, but among heterosexuals, males had higher proportions than females (31.5% vs. 19.7%). Divided assigned sex at birth, degree holders were the most prevalent among heterosexual males and homosexual females (42.8% and 49.3%) (**Table 2** & Supplementary Table 5 & **Figure 4**).

**Figure 4.**
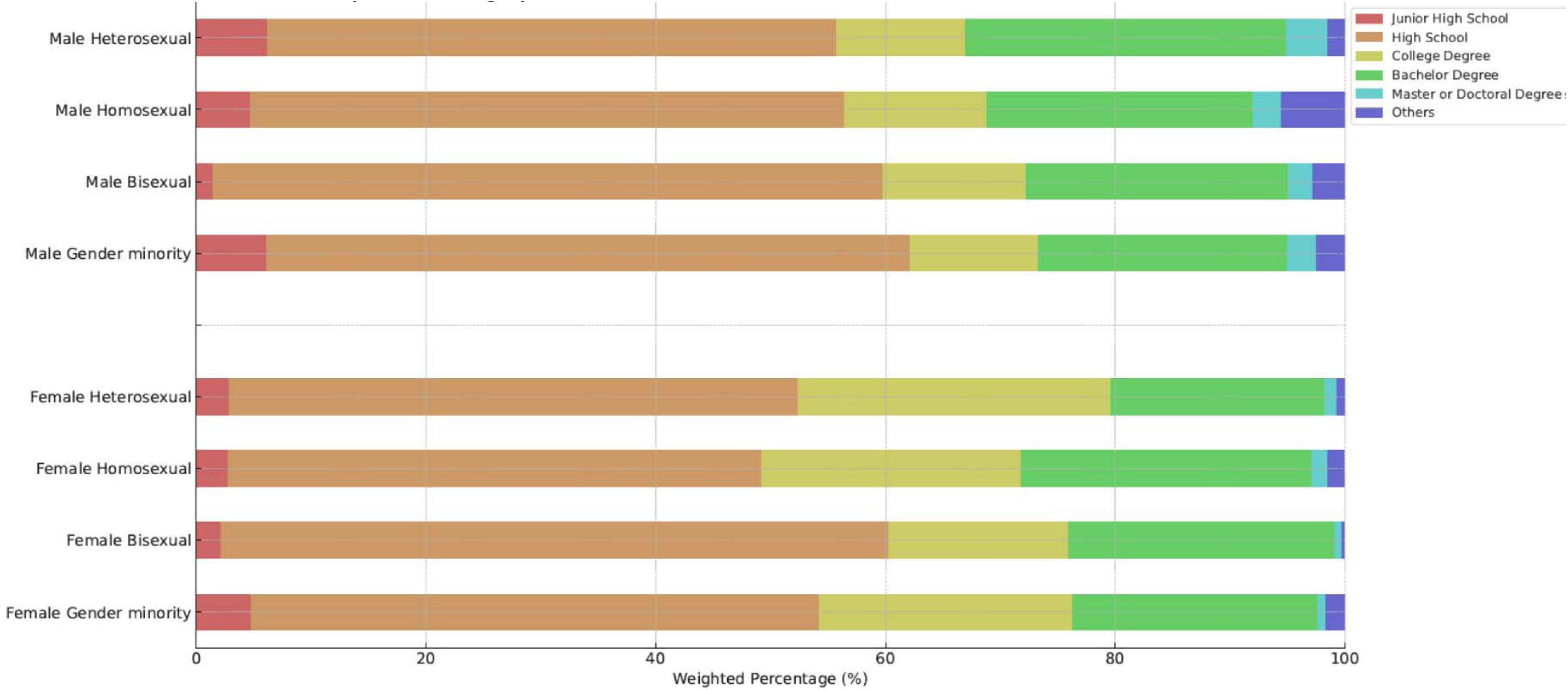
Academic attainment by SOGI category and sex. Abbreviation: SOGI, sexual orientation and gender identity

Employees/employed/self-employed was higher among homosexuals and bisexuals than heterosexuals (67.8%, 67.3% vs. 64.3%), with GMs having the lowest rates (57.2%). Heterosexual, homosexual, and bisexual males showed a decreasing and increasing trend in the proportion for employee/employed/self-employed and for unemployed, respectively. On the other hand, the opposite trend was seen for females. GMs had the lowest employment rates (27.4% males, 21.1% females) (**Table 2 & Supplementary Table 6, Figure 5**).

**Figure 5.**
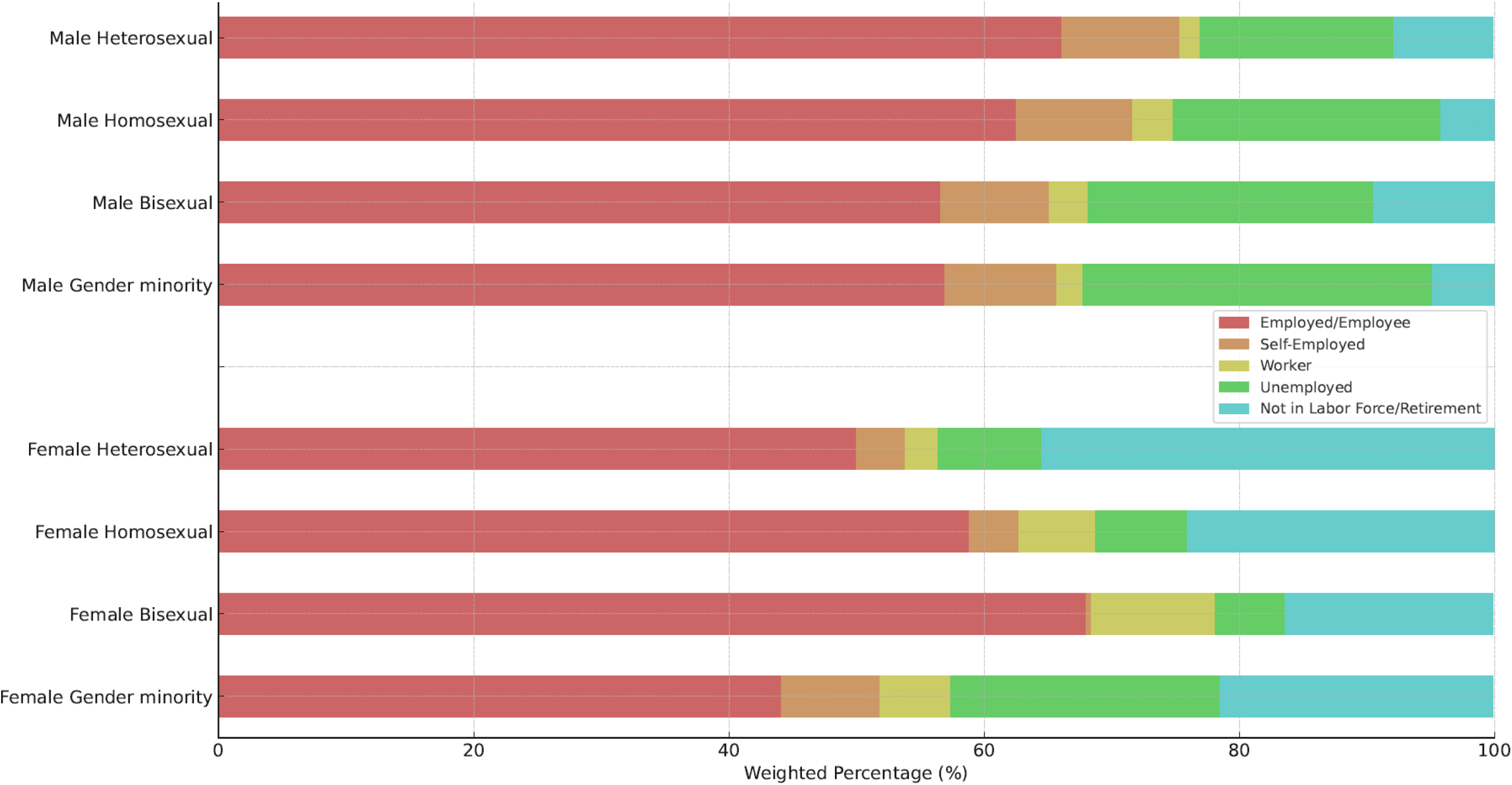
Employment status by SOGI category and sex. Abbreviation: SOGI, sexual orientation and gender identity

SGMs had lower health insurance coverage than heterosexuals (96.1% heterosexuals, 87.4% homosexuals, 89.4% bisexuals, 86.3% GMs), with males having less coverage than females in all SOGI categories. (**Table 2 & Supplementary Table 7, Figure 6**).

**Figure 6.**
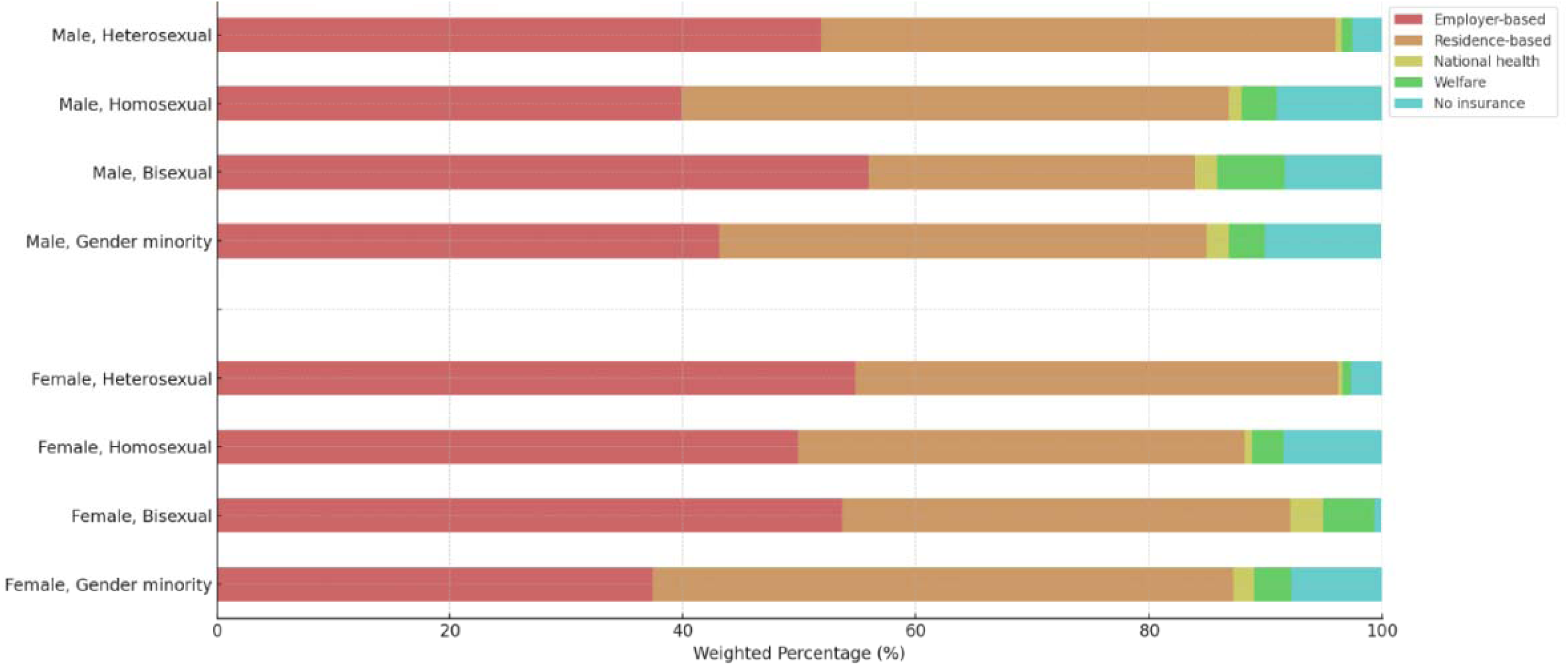
Health insurance coverage by SOGI category and sex. Abbreviation: SOGI, sexual orientation and gender identity

**Figure 7.**
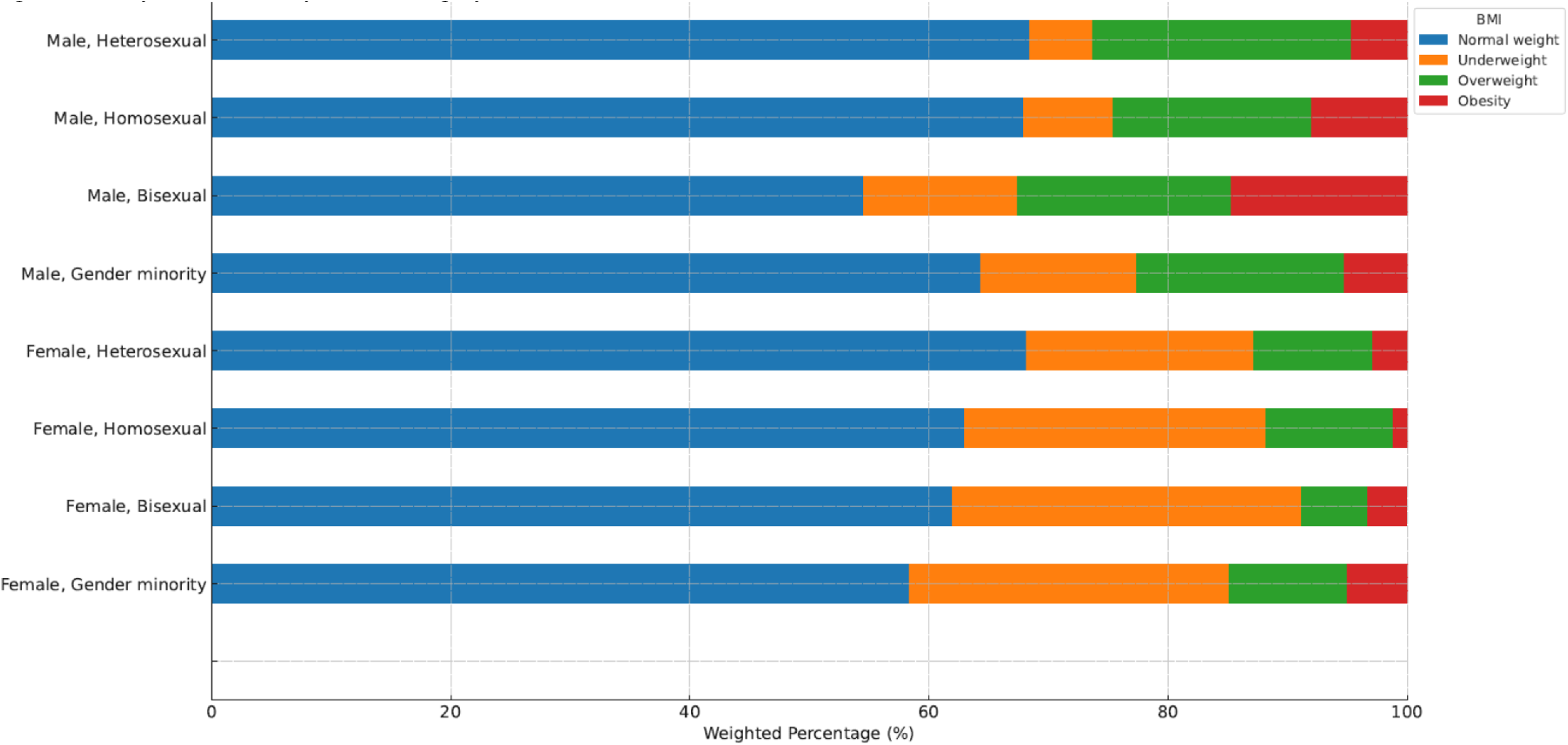
Body mass index by SOGI category and sex. Abbreviation: BMI, body mass index: SOGI, sexual orientation and gender identity

**Figure 8.**
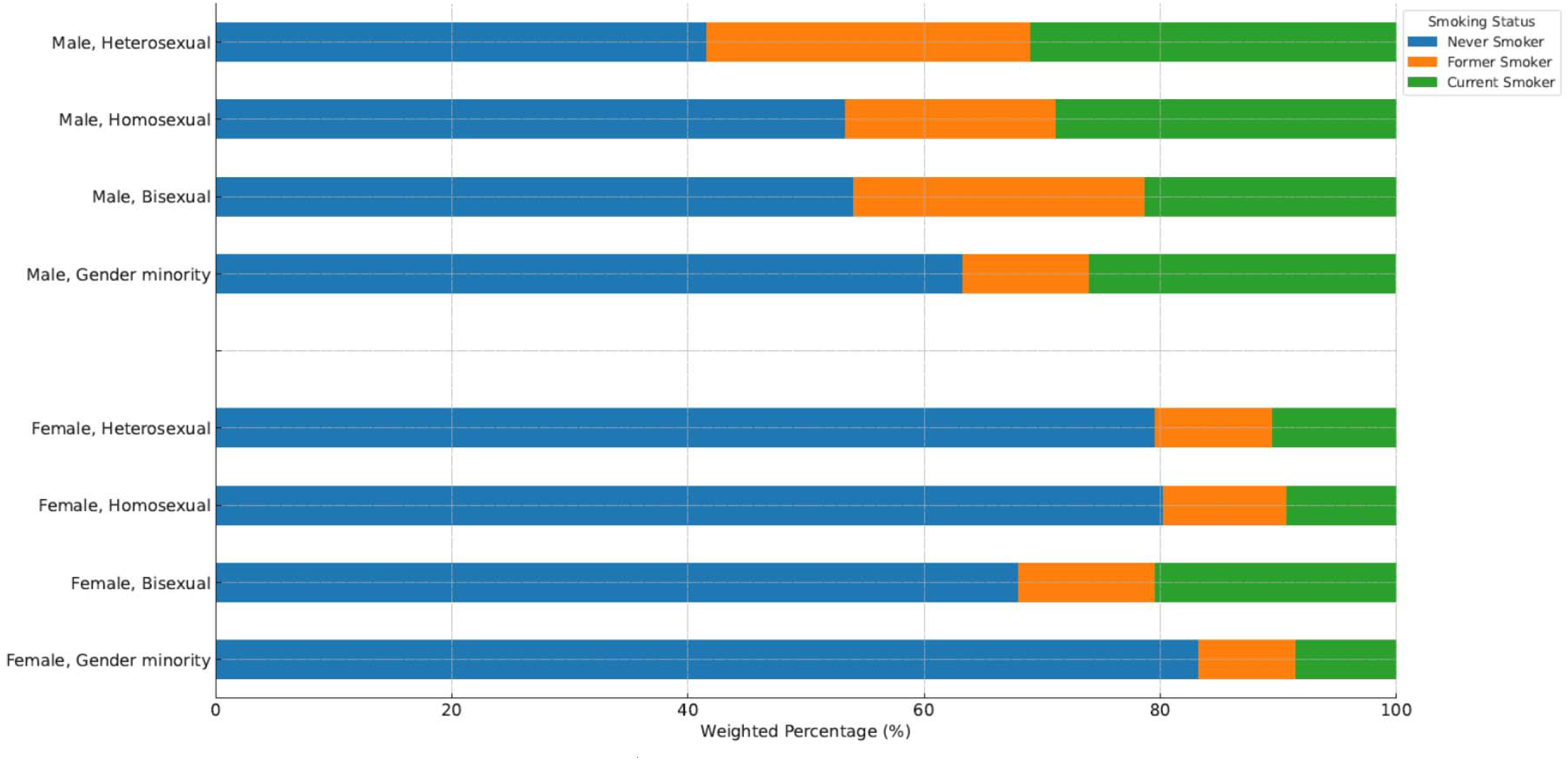
Smoking status by SOGI category and sex. Abbreviation: SOGI, sexual orientation and gender identity

Home ownership was lower among SGMs than heterosexuals (74.9% heterosexuals, 68.5% homosexuals, 70.4% bisexuals, 70.3% GMs), with bisexual males and homosexual females having the lowest among males and females, respectively (64% and 66.6%) (Table 2 **& Supplementary Table 8**).

The weighted average BMI was about 22 for all SOGI categories, with females less overweight/obese and more underweight than males. Homosexual and bisexual males were more likely to be obese and underweight than heterosexuals (8.0%, 14.7% obese vs. 4.7%; 7.5%, 12.9% underweight vs. 5.3%), and GMs were more likely to be underweight (13.1% vs. 5.3%). SGM females were more underweight than heterosexuals (25.2%, 29.3%, 26.7% vs**. 19.0%) (Table 2, Supplementary Table 9 & Supplementary** Figure 1).

Fewer GMs were current smokers than other SOGI categories. Bisexual and GM males were less likely to smoke than heterosexuals (21.3%, 25.9% vs. 31.0%), while bisexual females were more likely to smoke than others (20.4% vs. 31.0% heterosexuals, 9.3% homosexuals, 8.5% GMs) (Table 2 & **Supplementary Table10** & **Supplementary** Figure 2).

Fewer SGMs were habitual drinkers compared to heterosexuals, with a similar trend in males (40.6%, 35.2%, 34.4% vs. 51.7%). Among females, the trend was less pronounced (21.3%, 19.6%, 23.1% vs. 25.3%). Substance use was slightly higher among SGMs than heterosexuals (Table 2 & **Supplementary Table 11**).

SGMs had more medical comorbidities, including pneumonia/bronchitis, caries, COPD, chronic hepatitis/cirrhosis, immunodeficiency diseases, and psychiatric diseases. Allergic disease, cardiovascular disease, chronic renal failure, immunodeficiency disease, and cancer were more prevalent among SGM males (Table 2 & **Supplementary Table 12**).

SGMs reported lower self-rated health and higher psychological distress than heterosexuals. Bisexuals and GMs scored higher on the UCLA Loneliness Scale than heterosexuals and homosexuals. On the Fear of COVID-19 Scale, homosexual and GM males scored higher than heterosexual males (18.2, 18.8 vs. 16.9). Homosexual females scored higher than heterosexual females (19.4 vs. 17.8). SGMs were less likely to be fully vaccinated or boosted against COVID-19 compared to heterosexuals (males: 79.3% homosexuals, 81.4% bisexuals, 81.2% GMs vs. 88.0%; females: 83.3%, 80.9%, 85.4% vs. 87.3%) (Table 2 & **Supplementary Table 13)**

## Discussion

This is the first study aimed to estimate the proportion of SGM in Japan and describe their socioeconomic status, health-related conditions, and COVID-19-related variables using nationwide internet survey data. Our primary finding revealed that 4.8% of Japanese adults identified as homosexual, 1.3% as bisexual, and 3.8% as GM, providing nationally representative data. This indicates a significant portion of the Japanese adult population identifies as SGM, emphasizing the need to address their specific needs and concerns. Our study enhances the current literature by offering a more precise and representative estimate of the SGM population in Japan. Additionally, we found notable differences in socioeconomic and health-related status between SGM individuals and their heterosexual counterparts. Despite Japan’s relatively accepting cultural stance towards homosexuality, SGMs experienced partially worse socioeconomic and health-related outcomes.

The increased proportion of SGMs among younger generations, along with a higher proportion of homosexuals among men and a higher proportion of bisexuals and GMs among women, aligns with prior research conducted in the West.^18–22^

We found that a substantial proportion of Japanese SGMs, even homosexuals, were involved in opposite-sex marriages/partnerships. While in same-sex marriages/partnerships, homosexuals formed parenting households with comparable proportion to opposite-sex ones. The opposite-sex marriages/partnerships and parenting households in Japanese SGM may underlie “the familial norms of marriage and procreation” in Confucianism culture or may reflect the parallel between being in opposite-sex marriages/partnerships and conducting a same-sex relationship.^7^

On the other hand, there is scant research detailing opposite-sex marriages among SGMs. Similarly, mixed orientation marriages/relationships, namely those in a marriage/relationship whose sexual orientations do not match, are not well-studied to date^23^. The heterosexual marriages/relationships proportion of males who have sex with males was reported to be 3-9% in the US^24,25^, 11-60% in India^26^, and 33-51% in China.^27^ In contrast, those of females with sex with females were unclear despite qualitative studies encompassing limited participants.^28,29^ Such studies were less generalizable due to the sampling bias of the study participants, although the proportion of mixed-orientation marriages/relationships was higher in Asian countries compared to the West. Some studies revealed that SGM males in a mixed orientation marriage suffered from relationship distress^30^ or less satisfaction than in a heterosexual marriage^31^; meanwhile, those in China were associated with fewer anxiety symptoms than single.^10^ These inconsistent results could be attributed to the diverse norms of each country.

Concerning the association between income and SOGI category, this study was comparable to existing studies. Although there was variability in the reported income indicators, in general, SGMs were reported to have lower incomes, with bisexual women reported to have the lowest incomes, mainly based on Western studies.^19,20,32^ In addition, females had lower incomes than males in each SOGI category. Furthermore, in the study that included the ‘something else’ category (equivalent to GM in this study) ^20^, this category was reported to have the lowest income of all SOGI categories.

This study demonstrated disparities in academic attainment between SOGI categories in Japan and different distributions between Japan and other countries. Existing studies have consistently indicated that homosexual and bisexual males exhibit higher educational achievement than heterosexual males.^19,20,32^ However, in the case of Japan, the opposite trend was observed among males. Conversely, among females, prior studies have shown inconsistent results, suggesting that homosexual and bisexual females tend to possess higher academic attainment than heterosexual females ^20^, or they were ranked in the order of homosexuals, heterosexuals, and bisexuals, similar to the results of this study ^19^. In Japan, our findings align with the latter pattern.

Previous studies have found that male and female SGMs were less likely to be employed ^18–20^; however, SGMs excluded from GM were more likely to be employed in Japan. This was mainly reflected in the high employment proportions of homosexual and bisexual females. GM males and females had the worst employment status, consistent with a previous study. ^20^

Male and female SGMs in Japan had a higher proportion for the uninsured than heterosexuals, although the numbers are minimal. This result was consistent with US studies ^18,19^, which do not apply to universal health coverage. The burden of access to healthcare due to uninsured in Japan could be a potential contributor to health inequalities between SGMs and heterosexuals.

It has been reported that SGMs were at a higher risk of housing instability, especially homelessness.^33^ However, regarding housing tenure, we showed that bisexual males and homosexual females were less likely to own a home than other SOGI categories. Nonetheless, substantial differences were not observed between the SOGI categories concerning housing instability, that is, freeload or temporary housing, which could be evaluated in this study.

The distribution of BMI among SGMs compared to heterosexuals was inconsistent with previous Western studies.^34,35^ Interestingly, this study showed the opposite of the association that homosexual and bisexual females tended to be overweight/obese ^34^, and SGM males were less likely to be overweight/obese.^35^

Curiously, the prevalence of current smokers and habitual drinkers was less in SGMs than in heterosexuals among each sex, unlike previous studies.^21,22^ The prevalence of substance users was greater in SGMs than in heterosexuals, as in previous Western studies ^36^, but at a very low prevalence.

Comorbidities among SGMs include immunodeficiency diseases such as HIV and HCV, as well as diabetes, asthma, respiratory disease, cardiovascular disease, cancer, and psychiatric disease have been reported to be related socioeconomic status, health care access, tobacco, and substance use.^37–39^ The disparity between heterosexuals and SGM regarding comorbidities in Japan might be explained in the same way.

Like this study, SGMs reportedly have poorer self-rated health, feel lonelier, and experience psychological distress than heterosexuals.^19,40^ Interestingly, no differences in loneliness were observed between homosexuals and heterosexuals in the description of this study, so future research with covariate adjustment is warranted.

The SGM population suffered more vulnerabilities due to the COVID-19 pandemic.^41^ In a study with Fear of COVID-19 as the outcome, SGM was significantly worse than heterosexuals ^42^, while vaccination coverage did not differ between SGM and heterosexuals. ^43^

This study has several limitations. Firstly, as it relies on self-reported data, results may be influenced by recall and social desirability biases. While prior SGM studies used trained interviewers to reduce such biases, web-based surveys are promising for investigating sensitive behaviors like SOGI.^44^ Comparison with interview-based surveys is needed.

Secondly, the inverse probability weighting method may overestimate attributes with few respondents, such as minors or older females.^45^ It’s important to assess if the covariates used for sampling weights ensure national representativeness. Further studies with other cohorts or surveys are needed for better national representativeness. Thirdly, the SOGI category classification might be inaccurate.^16^ß While this study classified SOGI by sexual attraction and respondents’ gender, future surveys should also ask about sexual expressions and behaviors. Directed questions in the SOGI questionnaire might exclude “questioning,” “asexual,” etc., underestimating GM. Finally, web-based surveys include variables unsuitable for questionnaires (e.g., asymptomatic comorbidities) or that cannot be measured (e.g., homelessness).

## Conclusion

We estimated the percentage of SGMs in the Japanese population and described and compared several demographic variables between heterosexuals and SGMs. We clarified the differences in socioeconomic status and health status, then we also found that marital status, academic attainment, employment status, BMI, and smoking and drinking status differ from those in previous Western studies. The results of our descriptive epidemiological study will motivate policy makers, health care providers, and researchers to generate, analyze, and evaluate hypotheses in SGM research in Japan. We also hope that the results will contribute to an important step towards ensuring the well-being of SGM and defending their rights in a society that respects diversity and promotes equality.

## Research in Context Panel

### Evidence before this study

Previous research on sexual and gender minorities (SGMs) has primarily focused on Western populations, revealing significant health disparities, socioeconomic challenges, and varying degrees of acceptance and discrimination. Limited studies have been conducted in Asia, particularly in Japan, where cultural, philosophical, and religious contexts differ. Existing data are often not nationally representative, and there is a lack of comprehensive understanding of the demographic, socioeconomic, and health-related status of SGMs in Japan. No prior studies have utilized a nationally representative sample to examine these aspects comprehensively.

### Added value of this study

This study is the first to provide a nationally representative estimate of the proportion of SGMs in the Japanese adult population, along with a detailed description of their demographic, socioeconomic, and health-related variables. By using a large-scale internet survey and applying inverse probability weighting for national representativeness, this study offers a more accurate and comprehensive understanding of the SGM population in Japan. It highlights differences in socioeconomic and health-related outcomes between SGMs and heterosexual individuals, contributing valuable data to the limited body of research on SGMs in non-Western contexts.

### Implications of all the available evidence

The findings underscore the necessity for targeted health policies and interventions to address the unique needs and disparities faced by SGMs in Japan. The study’s results suggest that while there are similarities between the challenges faced by SGMs in Japan and those in Western countries, there are also distinct differences shaped by cultural and societal factors. This highlights the importance of developing culturally appropriate policies and interventions rather than directly applying Western models. Policymakers, healthcare providers, and researchers should consider these findings to promote equality, improve health outcomes, and ensure the well-being of SGMs in Japan. Future research should continue to build on this study to further understand the dynamics of SGMs in Japan and other non-Western contexts.

## Data Sharing Statement

The original data can be shared at a reasonable request to the corresponding author for research and academic propose.

## Acknowledgments

No conflicts of interest are disclosed.

