## supplement for "Estimation of Sexual and Gender Minorities in the adult population of Japan: Descriptive Epidemiological Study utilizing a Nationwide Cross-Sectional Internet Survey"

**Supplementary methods**

This study was a nationwide cross-sectional internet survey that utilized data from the 3rd wave of the COVID-19 and Society Internet Survey in Japan (JACSIS 2022). The self-reported web-based survey was conducted from September 12th to October 19th, 2022, by Rakuten Insight Inc., a reputable internet survey agency.

Participants were recruited from a large survey panel managed by a major, nationwide internet research agency, Rakuten Insight (former Rakuten Research), which maintains a pool of 4.1 million panelists with Japanese nationality covering all social categories, such as education, housing tenure, and marital status, defined by the census in Japan {Rakuten, #1407}. The survey panel consisted of people recruited initially through services managed by the Rakuten agency group. At the time of registration, participants were required to provide information, such as sex, age, occupation, and area of residence, and to agree that they would participate in different research surveys with web-based written consent. Minors provided their consent with approval from their parents or guardians {Tabuchi, 2019 #1355}.

In the 1st wave of the JACSIS study (JACSIS 2020), we employed a multistage sampling methodology, utilizing prefectures as the primary sampling unit and stratifying into 14 strata by 10-year age (15-19, 20-29,…60-69, 70- years) and registered sex (male or female) as the secondary sampling unit. The sample was sourced from a population of 2.3 million registered users in 2019, randomly selected via email invitations. Data on the target population was collected until the pre-determined sample size of 28,000 was reached. {Miyawaki, 2021 #1408} A target sample size of 32,000 was predetermined in the JACSIS 2022. To implement the longitudinal study design, priority e-mail invitations were sent to respondents who participated in the JACSIS and the sister survey study (JASTIS) {Tabuchi, 2019 #1355}, resulting in 41,626 respondents being invited 27,329 individuals longitudinal responded (response rate was 65.7％).and 4,631 respondents from a pool of 4.1 million registered users in 2022 were recruited until the target sample size was reached for each stratum. The respondents were incentivized with "E-points," credit points for internet shopping on the Rakuten homepage.

The JACSIS study series is an annual longitudinal internet survey where the questionnaire is incorporated into the survey as the research question proposal is approved by the committee meeting for each wave of the study. In this study, the author proposed a descriptive epidemiological study of Japanese SGM as the research question and was approved to incorporate the developed SOGI questionnaire. It was the same questionnaire as a national comprehensive survey described below, or approved questionnaires from other researchers among the JACSIS committee were also applied to this descriptive epidemiological study {Tabuchi, 2024 #1409}.

**Supplementary tables and figures**

**Supplementary Table 1:**

**Supplementary Table 2:**

**Supplementary Table 3:**

**Supplementary Table 4:**

**Supplementary Table 5: Completed TRIPOD Checklist for Model Validation**

**Supplementary Table 6:**

**Supplementary Table 7.**

**Supplementary Figure 1: Flow-Chart of Participants Included In The Study**

**PsyMetRiC-HK**

Total Sample

*n* = 608

Baseline Age ≥16 and ≤35

*n* = 429

With Psychosis-Spectrum Disorder Diagnoses

*n* = 429

Follow-up between 1-12 years

*n* = 429

Without Metabolic Syndrome at Baseline

*n* = 416
